# Efficacy and Safety of Vitamin D for COVID-19: A Living Evidence Synthesis Baseline report

**DOI:** 10.1101/2025.10.24.25338314

**Authors:** Trinidad Sabalete, Juan Antonio Blasco, Rocío Rodríguez, Maria Piedad Rosario, María Ximena Rojas-Reyes, The Living Evidence to Inform Health Decision Research Group (LE-IHD)

## Abstract

**Objective:** This living systematic review aims to provide a timely, rigorous, and continuously updated summary of the available evidence on the effectiveness and safety of using high dose (>4.000 IU) of Vitamin D for the treatment of mild, moderate, or severe COVID-19, as well as in the prevention of progression to long COVID.

**Methods:** We are conducting a Living Evidence Synthesis (LES) following the Living Evidence to Inform Health Decisions (LE-IHD) framework. A stepwise approach was used for evidence identification, beginning with the identification of existing systematic reviews to build an evidence matrix of relevant primary studies. We then searched for additional primary studies published after the latest search dates of the included reviews. The Epistemonikos L·OVE (Living OVerview of Evidence) platform supports screening and selection, assisted by automated classifiers. Regular automated searches are conducted across major databases (e.g., Cochrane, MEDLINE, EMBASE, CINAHL, PsycINFO, and others), and ongoing trials are manually searched every three months in international trial registries. Two reviewers independently screen all results. We included randomized controlled trials assessing the effect of high dose (>4.000 IU) of Vitamin D use in patients with moderate to severe COVID-19 on outcomes including all-cause mortality, COVID-19-related hospitalization, intensive care unit (ICU) admission, quality of life, adverse events associated with vitamin D use or hypervitaminosis D, length of hospital stay, and incidence of long COVID or post COVID condition. We assessed the risk of bias of included RCTs using RoB2 and applied the GRADE approach to assess the certainty of the evidence for the outcomes of interest. This is the baseline report of this Living Evidence Synthesis (LES), including studies identified up to 30^th^ November 2024. Based on its conclusions, we revisited the research question to determine whether it is suitable to continue in a “living” mode.

**Results:** We identified nine RCTs meeting our criteria for inclusion, all of them assessed the effect of high dose (>4.000 IU) of Vitamin D compared to no treatment (standard of care) or placebo. Four RCTs with a total 1086 patients, used cholecalciferol as a single oral bolus of ≥ 100,000 IU and other four RCTs with a total 444 patients, used cholecalciferol in daily regimen (> 4,000 and < 100,000 IU daily doses for at least 7 days). Only one used calcifediol for ICU admitted COVID 19 patients. We present evidence independently for these two products.

Low certainty evidence suggests that the use of ≥ 100,000 IU of cholecalciferol as a single bolus may increase all-cause mortality (RR=1.44, 95%CI: 0.88−2.37) and resulted in no differences regarding ICU admission (RR 0.93, 95 % CI 0.70−1.23; 11 fewer per 1000 patients, 95 % CI from 47 fewer to 36 more), and little to no difference in length of hospital stay. Three studies with 538 patients provide very low certainty evidence that suggest little to no effect of ≥ 100,000 IU of cholecalciferol on adverse events (4 fewer per 1000 patients, 95 % CI from 69 fewer to 186 more; RR 0.96; 95 % CI 0.33-2.81).

Low certainty of evidence suggests that *cholecalciferol in daily regimen* (> 4000 and < 100000 IU daily doses, for >7 days) may reduce all-cause mortality (2 studies; 265 patients; RR 0.79, 95 % CI; 0.60 to 1.03) and may results in little to no difference in length of hospital stay and adverse events compared to not use Vitamin D. The evidence is very uncertain about the effect of *cholecalciferol in daily regimen* on ICU admission (3 studies; 315 patients; RR 0.30, 95 % CI 0.08−1.18).

Finally, the evidence about the use of *calcifediol* in COVID -19 patients is very uncertain about its effect on all outcomes of interest.

**Conclusions:** Current available evidence does not support the use of vitamin D supplements in patients with COVID-19. The evidence for either benefit or harm from vitamin D supplementation is of low to very low certainty, with no significant differences observed between the various interventions (bolus vs. daily regimen) or between the products assessed (cholecalciferol and calcifediol).

Several ongoing trials may provide additional evidence to improve the certainty regarding the effects of vitamin D at varying dosages. This justifies the adoption of a Living Evidence approach to continuously monitor and update these conclusions as new data emerge.

We will keep this review in a “living mode” until either (a) the certainty of the evidence for benefits and harms becomes moderate to high, or (b) 24 months of surveillance are completed, whichever comes first.

The ongoing review focuses on high-dose vitamin D supplementation (>4,000 IU/day) for the prevention and treatment of severe COVID-19, including outcomes such as hospitalization, ICU admission, quality of life, all-cause mortality, and adverse events in patients with mild, moderate, or severe illness.

## BACKGROUND

The role of vitamin D in the prevention and/or treatment of COVID-19 is based on a potential immunomodulatory effect of innate and adaptative immune responses. Vitamin D receptor is expressed on immune cells (B cells, T cells and Ag-presenting cells), and these cells can synthesize the active vitamin D metabolite (1,2).

Recently studies in the UK Biobank cohort identified through National Health Service registers, found no independent associations between 25(OH)-vitamin D status and risk of severe infection or mortality for COVID-19, suggesting the presence of confounding factors (3,4).

However, other studies show a negative correlation between vitamin D levels or sunlight exposure and cases of COVID-19 in several countries (5,6).

Also, the relationship between vitamin D and COVID-19 was examined in the Spanish population identified through registers. A study analysed the results of vitamin D supplementation in COVID-19 outcomes, using the database of the public healthcare system in Catalonia (7). Another study analysed the data of Andalusian patients of the Public Health System with COVID-19 through their electronic health records in the Population Health Base (8). Both studies showed a reduction in mortality associated with vitamin D prescription in hospitalised patients with COVID-19.

A recent systematic review (SR) reported low certainty of the effects of high-dose of vitamin D for all-cause mortality, intensive care unit admission, and likely results in little to no difference in length of hospital stays (9). A wide variability of SRs currently published was observed (10–34).

The potential availability of new studies as several ongoing studies have been identified, we proposed to carry out a living SR.

This approach will make it possible to develop a rigorous and updated synthesis of the evidence on all-cause mortality, COVID-19-related hospitalisation, intensive care unit admission, quality of life, adverse events associated with the use of vitamin D or hypervitaminosis D, length of hospital stay, and long COVID-19 rates and draw conclusions on the prevention of severe COVID-19 and/or treatment in patients with mild, moderate, or severe illness for COVID-19.

This base line report of the living evidence synthesis (LES) has been used to inform a rapid response for health decisions in Spain. and is being developed as part of the LE to Inform Health Decisions program [35], which supports health system organizations in the implementation of a living process for the development of the synthesis of the evidence to inform health decisions. The continuous update resulting from evidence surveillance will be available at the program’s website (https://livingevidenceihd.com/lesrepo/).

### Protocol and registration

This manuscript complies with the ‘Preferred Reporting Items for Systematic reviews and Meta-Analyses’ (PRISMA) guidelines [36]. A generic protocol stating the methodology of LES to be conducted within the LE-IHD Program’s projects has been published elsewhere, and adapted for this evidence synthesis /SR to the specificities of the question and registered in OSF.

## METHODS

This is a baseline synthesis report of a Living Evidence Synthesis (LES) its design and planning followed the Living Evidence to Inform Health Decisions (LE-IHD) framework-based tool and its handbook (13) and complies with Methods for Planning and Reporting LES checklist proposed by Bendersky et al. (37). Inclusion criteria were SR and RCT designs, that assessed the use of vitamin D for the prevention or treatment of COVID-19. We restricted our review to high-dose greater than 4,000 IU. Outcomes of interest were all-cause mortality, COVID-19 related hospitalisation, ICU admission, quality of life and adverse events. We did not use the outcomes as an inclusion criterion during the selection process.

### Evidence identification, screening and selection

The Epistemonikos “Living OVerview of Evidence” (L.OVE) platform (38) was used as a technological enabler for evidence identification, screening, and selection. The authors worked with the team maintaining the L.OVE platform in devising the literature search aimed to identify initially SRs and RCTs. Searches were performed from inception in all data bases following the Epistemonikos procedures. No date, publication status, or language restriction were applied to the searches.

For this baseline synthesis, complementary searches were conducted in MEDLINE, Embase, and Cochrane CENTRAL to ensure the retrieval of potentially missed randomized controlled trials (RCTs). Additionally, an alert was activated in Evidence Alerts (McMaster University, Premium LiteratUre Service [PLUS™], https://plus.mcmaster.ca/COVID-19/). The complete search strategy is provided in Appendix 1. The last search was conducted in September 2023.

Search results were screened independently by two reviewers in two phases: first, by title and abstract; then, through full-text review of pre-selected references. Discrepancies were resolved through consensus or consultation with a third reviewer. We included health technology assessment reports, systematic reviews, and RCTs.

During full-text review, we excluded studies with fewer than 20 participants, RCTs evaluating vitamin D in combination with other vitamins or minerals, studies that did not mention the intervention of interest in the abstract, and preprints unless they reported results from a registered RCT.

### Data extraction and management

Using standardized forms, one reviewer extracted data from each included study. We collected information on study design, participant characteristics (including disease severity and age), eligibility criteria, and details of the intervention and comparator. Additional variables included dosage and treatment regimen, route of administration, follow-up duration, and baseline disease severity (mild, moderate, or severe). We also recorded the study’s funding source and any conflicts of interest disclosed by the investigators.

### Risk of bias assessment

We assessed the risk of bias using the AMSTAR II tool for systematic reviews [39] and the Cochrane RoB 2 tool for randomized controlled trials [40]. One reviewer conducted the assessments for all included studies.

### Measures of treatment effect

For dichotomous outcomes, we extracted event counts to calculate risk ratios (RRs) with 95% confidence intervals (CIs). For continuous outcomes, we extracted means and standard deviations (SDs) to estimate mean differences and their 95% CIs.

We considered the following baseline factors as potential confounders: age, comorbidities (including chronic obstructive pulmonary disease, cardiovascular disease, congestive heart failure, diabetes, obesity, hypertension, chronic kidney disease, hyperlipidemia, immunosuppression, and cancer), co-interventions, and disease severity.

### Data synthesis

Given the fundamental differences between cholecalciferol and calcifediol, we analyzed these interventions separately. Among studies using cholecalciferol, we identified two subgroups based on dosage regimen: RCTs administering a single bolus ≥100,000 IU, and RCTs using doses <100,000 IU over more than one week. We summarized the characteristics of individual studies narratively.

We conducted meta-analyses using RevMan 5, applying the inverse variance method with a random-effects model. For outcomes lacking sufficient data for effect estimation, or when study heterogeneity precluded pooling, we reported findings narratively.

### Assessment of certainty of evidence

We assessed the certainty of the evidence for all outcomes using the Grading of Recommendations Assessment, Development and Evaluation working group methodology (GRADE Working Group) approach (41), across the domains of risk of bias, consistency, directness, precision and reporting bias. Certainty was adjudicated as high, moderate, low or very low.

We planned to identify significant differences between subgroups (test for interaction <0.05). In case of significant differences by subgroups we considered reporting the results of subgroups separately and if subgroup estimates including null effect.

We planned to perform sensitivity analysis excluding high risk of bias studies. In cases where the primary analysis effect estimates and the sensitivity analysis effect estimates significantly differed, we considered presenting either the low risk of bias — adjusted sensitivity analysis estimates.

For the main comparisons and outcomes, we prepared a Summary of Findings (SoF) table (42,43). Also, we used GRADEpro GDT software for developing an evidence profile for the main outcomes (44).

### Living evidence maintenance

We will keep this question in a living mode until the certainty of the evidence on the updated estimates for benefits and harms becomes moderate to high or after the next 24 months of surveillance, whatever is reached first. We will continuously monitor the evidence by performing daily searches and screening of the retrieved references each two months.

All potentially eligible studies will undergo the selection process described above as in the review protocol (9). Additionally, every three months, we will manually search for ongoing studies in the WHO International Clinical Trials Registry Platform (ICTRP) and the clinicaltrials.gov. If a potentially eligible study is found, a second reviewer will confirm its eligibility by reading the full test. All new eligible studies will undergo a data extraction process. The data synthesis will be updated immediately after that considering the predefined subgroups of interest. We will evaluate the statistical heterogeneity among included studies by using the I2 statistics. If new heterogeneity is detected (i.e. increase the heterogeneity previously identified or new heterogeneity arises where it was previously undetected), we will explore its potential sources by reviewing the new studies against previously included studies in order to identify reasons that may explain inconsistent results among studies. The body of evidence for the outcomes of interest will be assessed following the GRADE approach accordingly looking for changes in the certainty assessment results.

The living process will be supported by the LE-IHD Program. Results from evidence surveillance, along with all monitoring activities, will be recorded and stored using the LE-IHD framework-based tool. Information on PRISMA will be updated accordingly. A living, web-based version of this review will be openly available through the LE-IHD Program repository [https://livingevidenceihd.com/lesrepo/]. The review will be resubmitted whenever the conclusions change or when substantial updates occur.

## RESULTS

### Search results

Up to 30/07/2023, the searches had retrieved 256 documents in the L·OVE platform for this question. Following a stepwise approach for identifying evidence, 26 SRs were initially selected as they met the inclusion criteria [9–34]. The evidence matrix created from these SRs included 670 primary studies, of which only seven RCTs met our inclusion criteria and were included for data extraction [45–53]. Complementary searches of randomized controlled trials (RCTs) identified 29 RCTs, from which only 18 were published after the last date search of the included SRs. After full-text assessment all of them were excluded for the following reasons: six studies were not RCTs, two included different populations than specified in the PICO question, four addressed different interventions, four used different comparators, and four did not report main outcomes. Additionally, two RCTs meeting our selection criteria were identified later through COVID-19 Evidence Alerts and were also included in the evidence synthesis [48, 50].

We finally included a total of nine RCTs, all of them reported on all-cause mortality, intensive care unit (ICU) admission, length of hospital stays, and adverse events. No studies reported data on COVID-19-related hospitalization, quality of life, or long COVID-19 rates.

We identified 24 ongoing studies. No RCTs or non-randomized studies (NRS) were identified regarding the prevention of severe COVID-19. A detailed list of included, excluded and ongoing studies, along with reasons for exclusion, is provided in Appendix 2.

**Figure 1.**
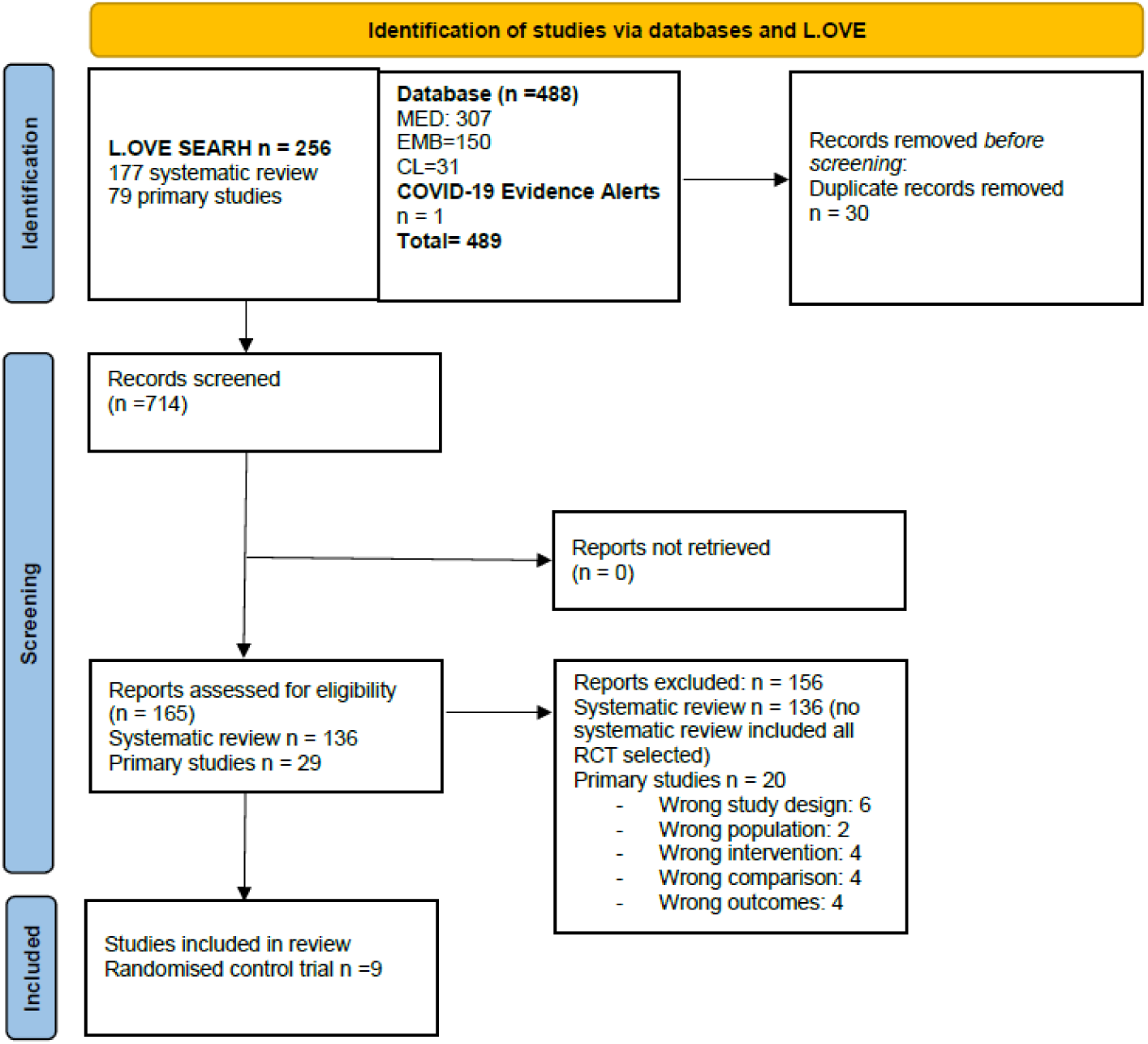
PRISMA Flowchart.

### Characteristics of included studies

A table summarising the characteristics of the included studies is included in the Appendix 1.

### Design of Studies and Setting

We included nine randomized controlled trials (RCTs) evaluating high-dose vitamin D (greater than 4,000 IU). Four studies used an open-label randomized design [46, 48, 49, 51], while five were double-blinded and placebo-controlled [45, 47, 50, 52, 53].

### Participants

All studies were conducted in hospitalized patients with COVID-19. Participants were randomly assigned to receive either vitamin D plus standard care or placebo/no intervention plus standard care.

A confirmed COVID-19 diagnosis by polymerase chain reaction (PCR) testing at enrolment was reported in eight studies. Cannata-Andía et al. assessed patients using either PCR or antigen testing [46]. Karonova et al. included patients with a positive PCR test and/or chest computed tomography (CT) [51]. Murai et al. confirmed SARS-CoV-2 infection using PCR or serological assay (ELISA) [53]. De Niet et al. did not specify the diagnostic procedures used [47].

### Interventions and Comparators

We categorized the studies into three subgroups according to key differences in the intervention:

- **High dose (> 4,000 IU) of vitamin D**: Cannata-Andía et al. administered 100,000 IU of cholecalciferol in a single oral dose at admission [46]. Jaun et al. used 140,000 IU as a single oral dose followed by 800 IU/day of vitamin D3 [50]. Mariani et al. administered a single 500,000 IU dose [52]. Murai et al. gave a single 200,000 IU dose [53].
- **Regimen (Extended-Duration) of Cholecalciferol** (<100,000 IU Daily): Bychinin et al. used 60,000 IU weekly, with daily maintenance of 5,000 IU, repeated on days 8, 16, 24, and 32, continuing until ICU discharge or death [45]. De Niet et al. gave 25,000 IU daily for 4 days, followed by 25,000 IU weekly for up to six weeks [47]. Domazet et al. administered 10,000 IU daily during ICU stay or for at least 14 days post-discharge [48]. Karonova et al. used 50,000 IU on days 1 and 8 of hospitalization [51].
- **Calcifediol Dosing:** Entrenas et al. used 0.532 mg of calcifediol on day 1, 0.266 mg on days 3 and 7, and weekly thereafter until discharge or ICU admission. The intervention was compared with placebo [49].

### Outcome Measures

The primary outcome, all-cause mortality, was assessed in seven studies [45, 46, 48–50, 52, 53]. ICU admission was reported in four studies [46, 50, 52, 53]. Length of hospital stay was assessed in eight studies [45–48, 50–53]. Adverse events were reported in six studies [45, 48, 50–53].

### Risk of bias of included SRs /primary studies

The overall risk of bias in the included studies was low. Six studies were assessed as low risk of bias and three studies with moderate risk of bias. The three studies with high risk of bias presented concerns in blinding of participants and personnel (performance bias) (46,49,51); two of these three studies had concerns in blinding of outcome assessment (detection bias) (46,51). Appendix 4 presents the traffic light figure and main results ofr this assessment.

### Effects of interventions

No randomized controlled trials (RCTs) or non-randomized studies (NRSs) meeting the criteria for the prevention of severe COVID-19 were identified.

For treatment effects in patients with mild, moderate, or severe COVID-19 illness, the included studies did not report on the following outcomes: COVID-19-related hospitalization, quality of life, and long COVID-19 rates.

Results for the outcomes all-cause mortality, intensive care unit (ICU) admission, length of hospital stay, and adverse events are presented below. A summary of findings, along with the GRADE evidence profile, is provided in Appendix 5.

### Comparison 1: High dose (> 4,000 IU) of vitamin D in patients with mild, moderate, or severe illness for COVID-19

#### 1. All-cause mortality (Figure 3; GRADE profile 1)

Four studies (n = 1086 participants) assessed the effect of *cholecalciferol* as a single bolus ≥ 100,000 IU on all-causes mortality (46,50,52,53). Low certainty evidence suggests that the use of *cholecalciferol* may result in an increase of all-cause mortality in COVID 19 patients (RR 1.44 (95 % CI 0.88−2.37) but the estimation is imprecise.

**Figure 3.**
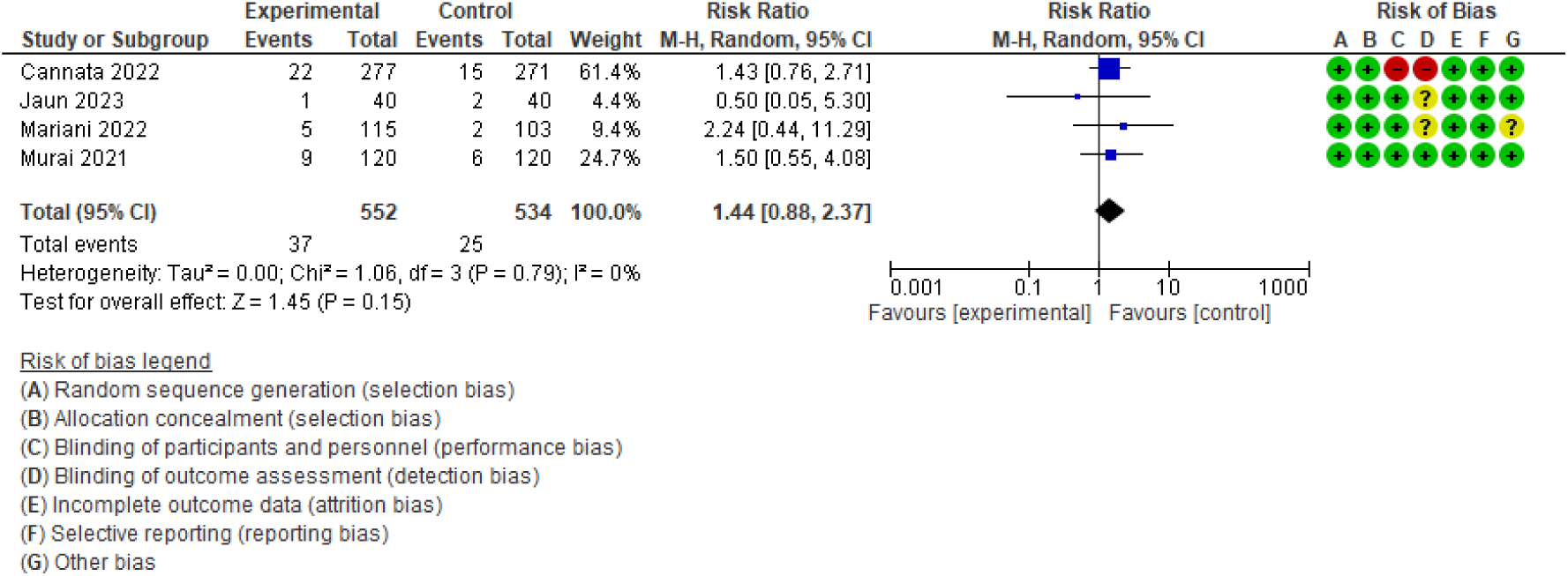
Forest plot for effect of cholecalciferol as a single oral bolus ≥ 100 000IU on all-cause mortality.

#### 2. Intensive care unit (ICU) admission

Four studies (n = 1086 participants) assessed the effect of *cholecalciferol* as a single bolus ≥ 100,000 IU on ICU admission (46,50,52,53). Low certainty evidence suggests the use of Vitamin D has little to no effect on ICU admission (RR 0.93; 95 % CI 0.70 to 1.23).

#### 3. Length of hospital stay

Four studies (n = 1006 participants) assessed the effect of ***cholecalciferol administered as a single high-dose bolus*** (≥100,000 IU) on length of hospital stay [46, 50, 52, 53]. However, the variability in effect measures across studies makes it unfeasible to meta-analyse their results. In one study, the median length of hospital stay was 6–7 days (IQR 4–10) in the intervention group versus 6–7 days (IQR 4–13) in the control group. In another study, the median hospital stay was 10 days (95% CI 9– 10.5) in the intervention group versus 9.5 days (95% CI 9–10.5) in the control group. A third study reported a median stay of 10 days in the intervention group versus 6 days in the control group, but no measures of dispersion or precision were provided, therefore we did not metanalyses these results.

The low-certainty evidence from unpolled data suggests little to no difference in the length of hospital stay between receiving or not receiving a single high-dose bolus of cholecalciferol.

#### 4. Adverse events associated with the use of vitamin D or hypervitaminosis D

Three studies (n = 538 participants) assessed the effect of ***cholecalciferol administered as a single high-dose bolus*** (≥100,000 IU) on adverse events (50,52,53). The evidence is very uncertain about of effect of cholecalciferol in single bolus on adverse events due to serious inconsistency and very serious imprecision (RR 0.96 (95 % CI 0.33-2.81), (figure 7). Among sources of heterogeneity, we explore variability in underlying diseases and intervention and any variability in the dose of cholecalciferol used. (GRADE profile – Appendix 5).

##### Comparison 2: Regimen o**f** at least 7days of vitamin D in patients with mild, moderate, or severe illness for COVID-19

#### 2.1. All-cause mortality (Figure 4; GRADE profile 1)

**Figure 4.**
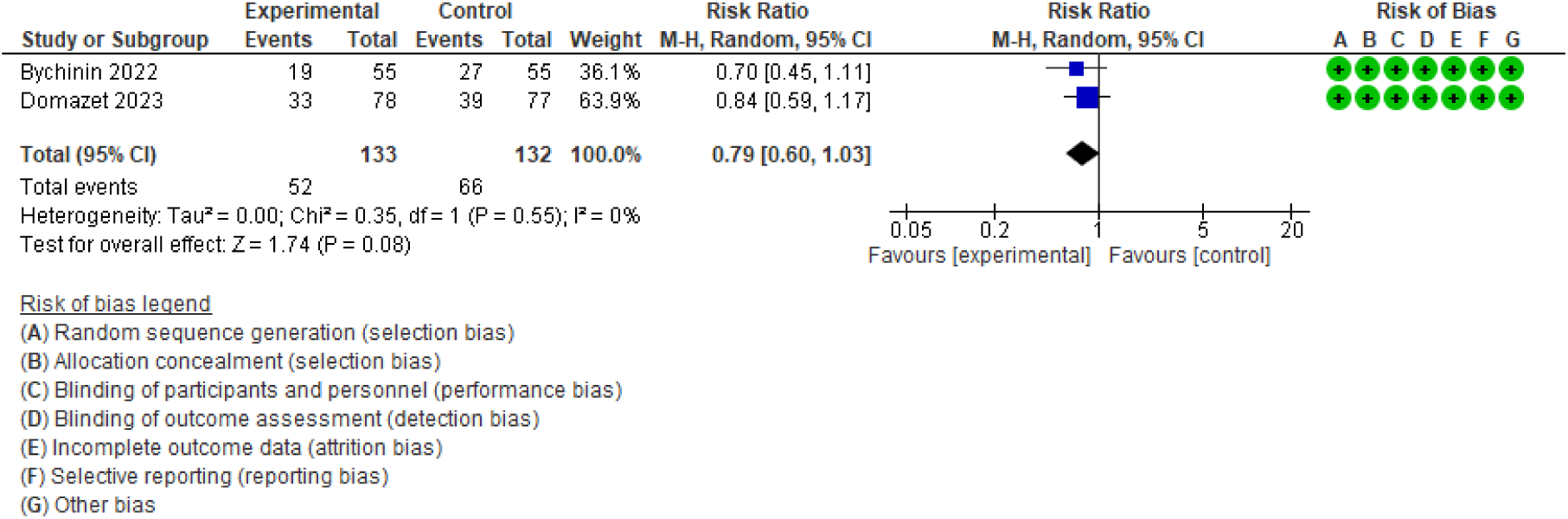
Forest plot of for effect of cholecalciferol treatment> 7days with dose/day > 4000 and < 100000) on all-cause mortality.

Two studies (n = 265 participants) assessed the effect of *cholecalciferol* regimen of at least 7days treatment with daily doses > 4000 and < 100000 (45,48) on all-cause mortality (45,48). Low certainty evidence suggest that this regimen of Vitamin D may result in a little reduction to no effect in all-causes mortality (RR 0.79; 95 % CI 0.60 to 1.03].

#### 2.2. Intensive care unit (ICU) admission

Two studies (n = 179 participants) assessed the effect of *cholecalciferol* regimen on ICU admission (47,51). We found RR 0.30, 95 % CI 0.08−1.18). (Figures 5 and 6). The evidence is very uncertain about of effect of *cholecalciferol* regimen on ICU admission due to serious risk of bias and very serious imprecision (very low certainty evidence). See GRADE evidence profile in Appendix.

**Figure 5.**
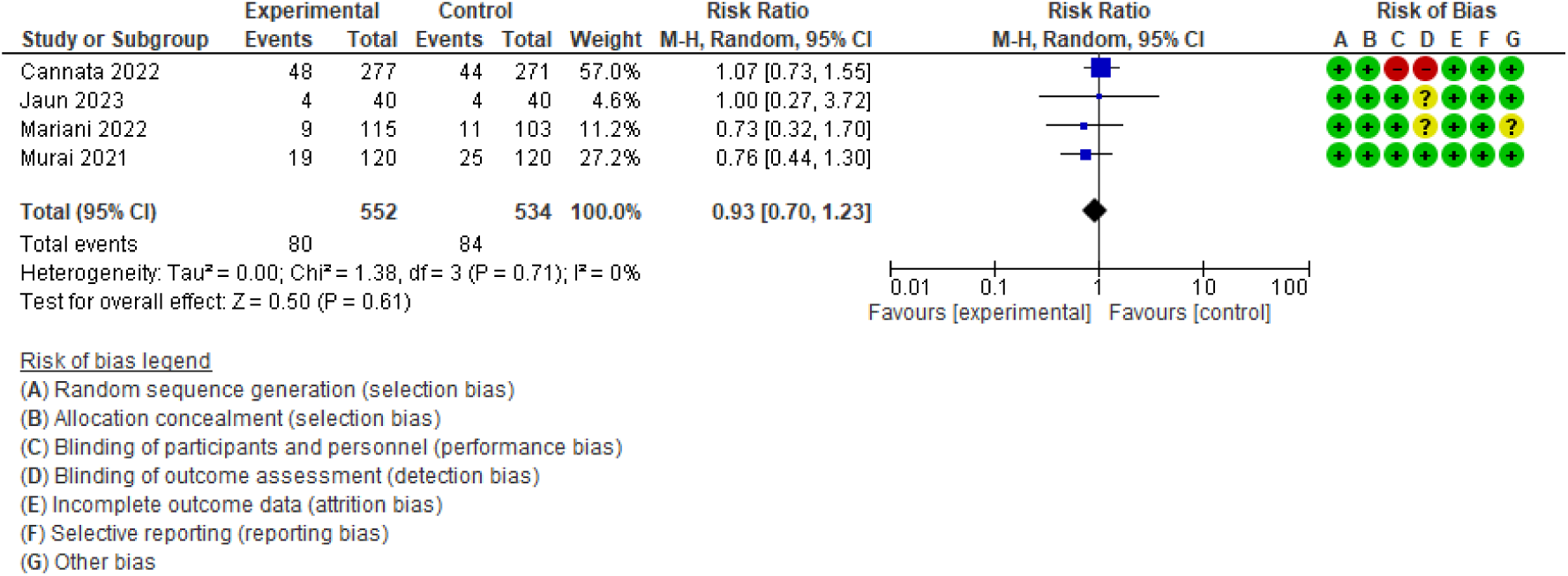
Effect of a single oral bolus ≥ 100 000IU of cholecalciferol on ICU admission.

**Figure 6.**
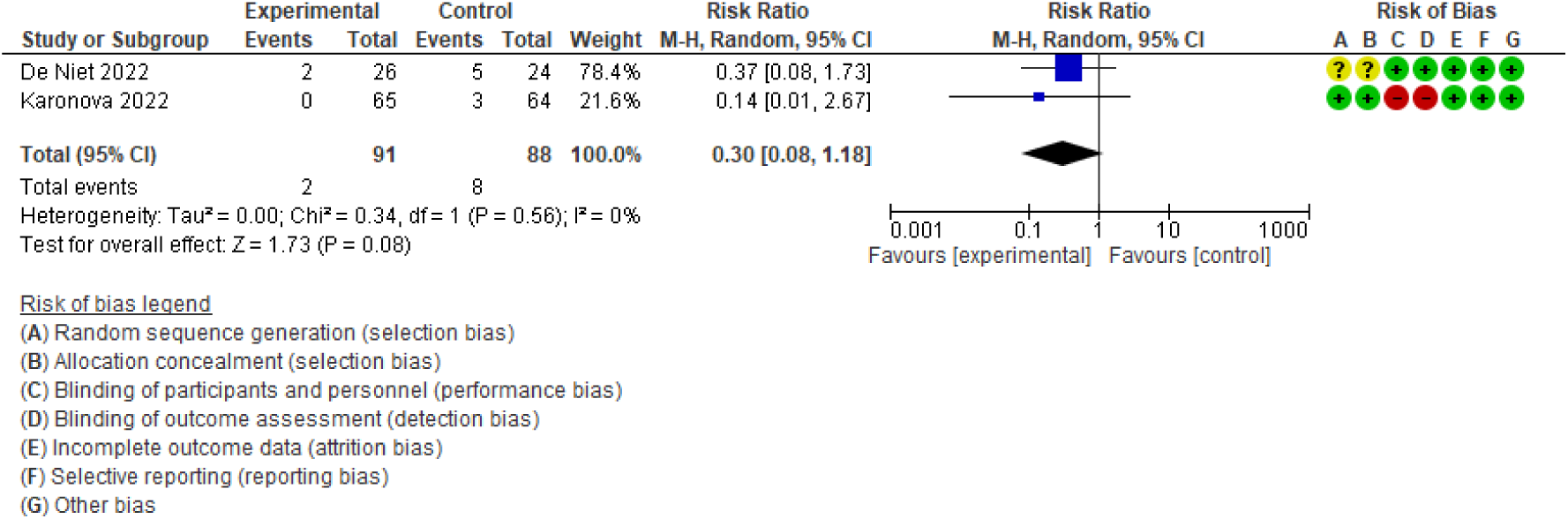
Effect of cholecalciferol regimen (> 7days with dose/day > 4000 and < 100 000 IU) on ICU admission.

**Figure 7.**
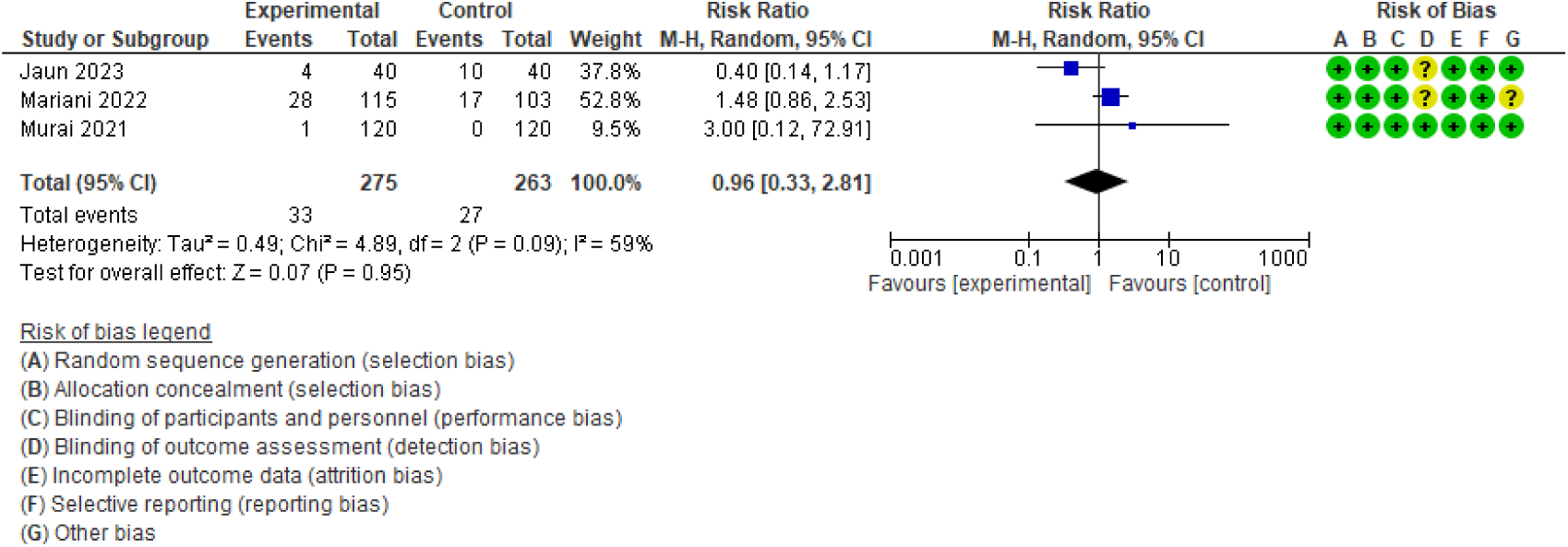
Effect of a single oral bolus ≥ 100 000IU of cholecalciferol on adverse events.

#### 2.3. Length of hospital stay

Four studies (n = 444 participants) assessed the effect of ***cholecalciferol regimen*** (≤4.000 < 100,000 daily doses for at least 7 days) on length of hospital stay (45,47,48,51). The median values in these studies were 15 days in intervention group (3-25.8 IQR) and 14.37 in control group (6-25 IQR). Low certainty evidence suggested a little to no difference in length of hospital stay.

#### 2.4. Adverse events associated with the use of vitamin D or hypervitaminosis D

Three studies (n = 315 participants) assessed the effect of ***cholecalciferol administered as a daily regimen*** on adverse events [45, 47, 48]. One additional study did not report any information on adverse events [49].

##### Comparison 3: Calcifediol dosing in patients with mild, moderate, or severe illness for COVID-19

###### 3.1. All-cause mortality (GRADE profile 1)

One study (n = 76 patients) assessed the effect *calcifediol* in all-cause mortality (49). The evidence is very uncertain about the effect of calcifediol on this outcome (RR 0.13; 95 % CI 0.01 to 2.73).

###### 3.2. Intensive care unit (ICU) admission

None of the studies assessed the ***ICU admission*** for calcifediol.

###### 3.3. Length of hospital stay

None of the studies assessed the ***length of hospital stay*** for calcifediol.

###### 3.4. Adverse events associated with the use of vitamin D or hypervitaminosis D

The adverse events associated with calcifediol was not evaluated in any study.

## Discussion

This baseline review, conducted as part of a Living Evidence Synthesis (LES), included nine randomized controlled trials (RCTs) evaluating the use of high doses of vitamin D (>4,000 IU/day) compared to placebo or no intervention for the prevention or treatment of COVID-19 in patients with mild, moderate, or severe illness. The results show that current evidence does not allow for firm conclusions about whether vitamin D improves outcomes in this population. Across all assessed outcomes, the evidence is limited by imprecision (e.g., wide confidence intervals and failure to meet the optimal information size), resulting in low or very low certainty.

The nine published RCTs are heterogeneous in terms of vitamin D delivery method and dosage, baseline disease severity, and participant risk factors—many of which were not consistently reported or available. Four studies (46, 50, 52, 53) assessed the effect of cholecalciferol administered as a single bolus, four studies (45, 47, 48, 51) evaluated a regimen of cholecalciferol given over more than seven days, and one study (9) assessed the effect of calcifediol in COVID-19 patients.

We considered three groups of interventions due to variability in form and dose of vitamin D assessed in the included studies: i) studies using cholecalciferol as a single bolus; ii) studies that using cholecalciferol regimen > 7 days, and iii) studies using calcifediol. Overall, the findings suggest no effect of cholecalciferol or calcifediol on key clinical outcomes.

Major limitations of this review are related to the completeness of the evidence. Despite reporting on critical outcomes such as all-cause mortality, ICU admission, adverse events, and length of stay, none of the studies reported on COVID-19-related hospitalization, quality of life, or long COVID-19 outcomes. All-cause mortality, COVID-19-related hospitalisation, ICU admission, quality of life, adverse events, length of hospital stay, and long COVID-19 rates resulted in insufficient power to draw conclusions.

Currently, considerable number of studies are still ongoing.

### Overall completeness and applicability of evidence

There was a lack of information regarding outcomes such adverse events in three of nine included studies.

We discarded Non-randomized studies (NRS) due to significant number of RCTs findings. We will evaluate in next versions for inclusion of NRS to analyse long-term adverse events.

Interventional trials are needed to uncover possible differences between calcifediol/calcitriol and cholecalciferol supplementation and to distinguish the effect of intervention in prevention and treatment.

### Comparison to other similar publications

In recent years, research in this field has shown important uncertainties about the beneficial effect of vitamin D on the prevention or treatment of COVID-19 and heterogeneity of interventions in the RCTs published (9).

### Limitations and strengths

This SR has a few strengths: We developed this review using a robust predefined methodology stated in a registered protocol. All steps were done in duplicate thus limiting errors throughout the review process. The protocol was planned to summarise the evidence using the GRADE methodology which is a highly transparent methodology for evidence assessment. It was not possible to pool the outcomes of all included studies for heterogeneity of interventions. The review was limited by the low numbers of events for some outcomes of interest combined with unmet optimal information size for multiple outcomes which made imprecision an important issue.

### Clinical implications

This review provides important evidence-based guidance regarding the role of high dose (> 4000 IU) of vitamin D in the prevention of severe COVID-19 and/or treatment in patients with mild, moderate, or severe illness for COVID-19.

Therefore, interventional trials are needed to uncover possible differences between calcifediol/calcitriol and cholecalciferol supplementation in critically ill COVID-19 patients.

### Implications for research

This evidence synthesis is part of a larger project set up to put the LE approach into practice. This project aims to produce multiple parallel living systematic reviews relevant to inform decisions, following the higher standards of quality in evidence synthesis production. We believe that our methods are well suited to handle the evidence that is to come, including evidence on the role of high dose >4,000 IU of vitamin D.

We have identified a number of ongoing studies addressing this question, including 24 RCTs, which will provide valuable evidence to inform researchers and decision makers in the near future.

During the next year, we will maintain a living, web-based, openly available version of this review at the Living Evidence to Inform Health Decisions website (https://livingevidenceframework.com/en/lesr/), and we will re-submit the review for publication every time the conclusions change or whenever there are substantial updates. Our SR aims to provide a high-quality, up-to-date synthesis of the evidence that is useful for clinicians and other decision makers.

## Data Availability

All data produced in the present study are available upon reasonable request to the authors

## Acknowledgments

We would like to acknowledge the contribution of the Living Evidence to Inform Health Decisions Program research group (Ariadna Auladel, Josefina Bendersky, Gerard Urrutia) to the development of this LES. The Epistemonikos Foundation team in charge of designed and conducted the searches and provided technological support in the L.OVE platform.

Also, we would like to acknowledge the per review of Soledad Isern, and the participation of Rebeca Isabel-Gómez in the initial process of developing the search strategies and subsequent updates in the databases: Medline, Embase and Cochrane Library.

## Roles and contributions

The idea was conceived by Juan Antonio Blasco, Rocío Rodríguez, María Piedad Rosario and Trinidad Sabalete. Rocío Rodríguez and Trinidad Sabalete conducted initial searches and wrote the first draft of the protocol, with significant contributions from all other authors. María Ximena Rojas provided methodological advice for the definition of the living evidence approach to be followed in this project, contributes with the GRADE certainty of evidence assessment and made important intellectual contribution to the final version of the review.All the authors and the LE-IHD program research group reviewed and approved the final version of the manuscript.

## OSF registration

*CRD42020181032*

## Ethic1s

As researchers will not access information that could lead to the identification of an individual participant, obtaining ethical approval was waived.

## Appendix 1 Boolean search strategy

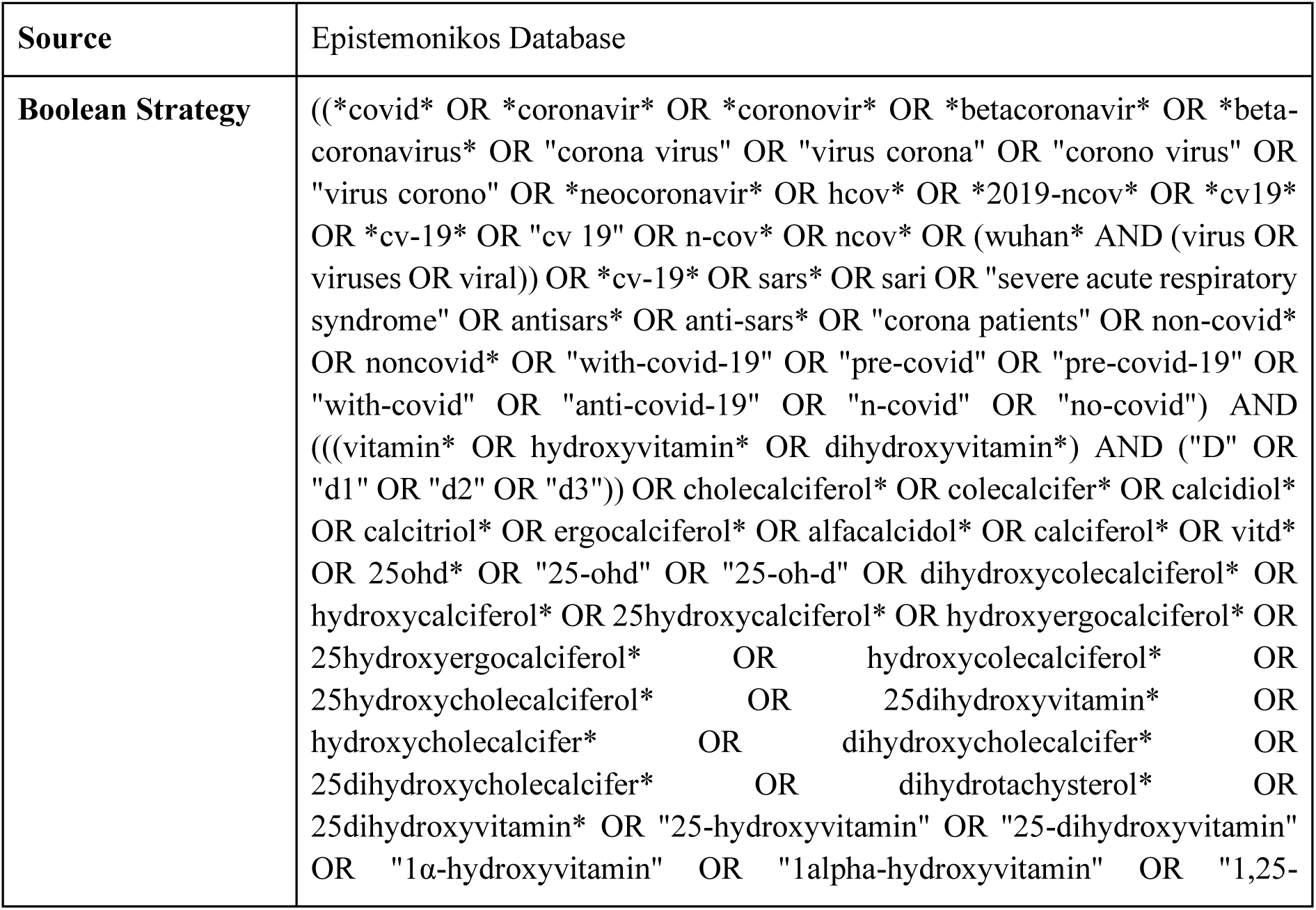
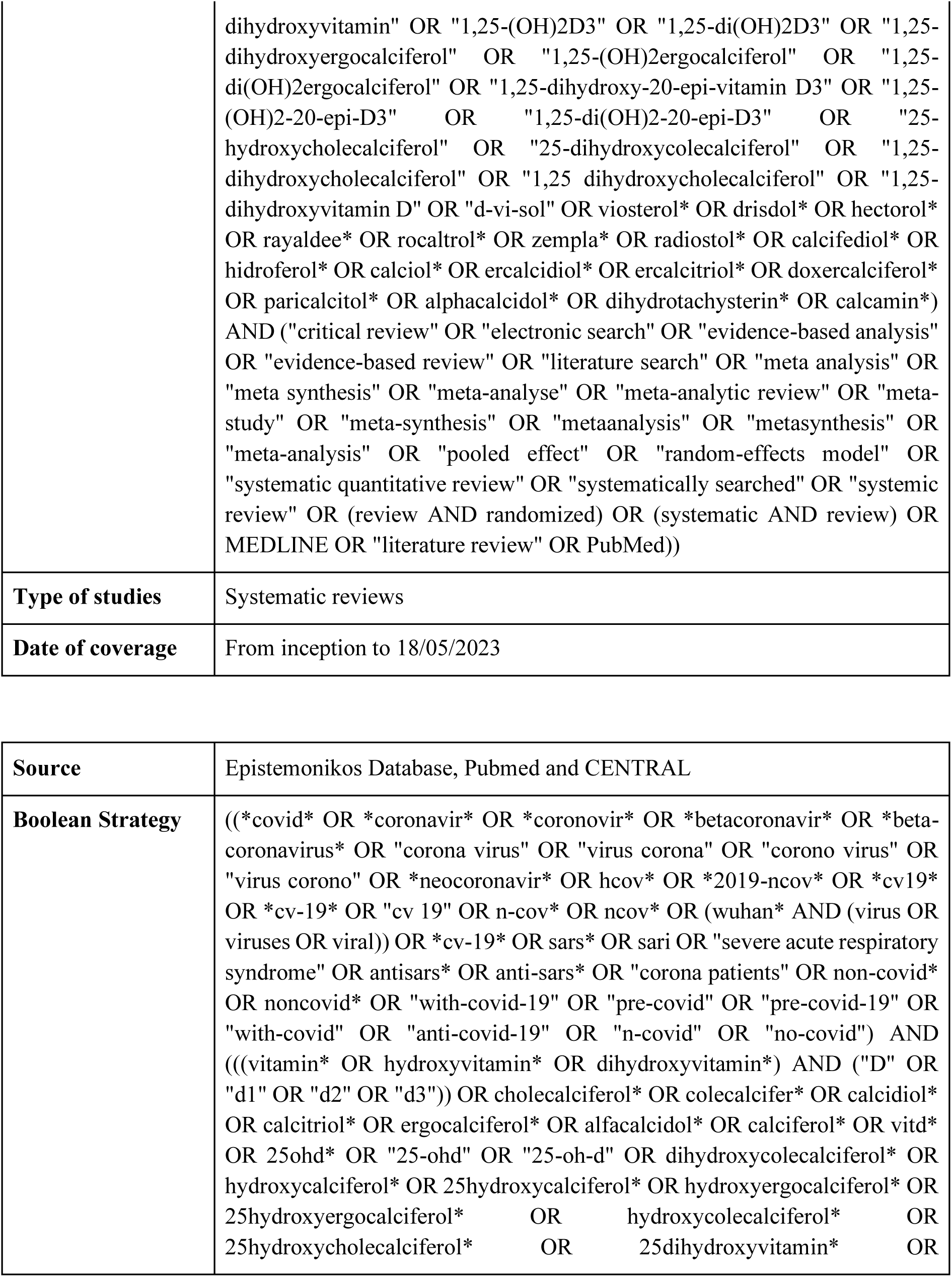
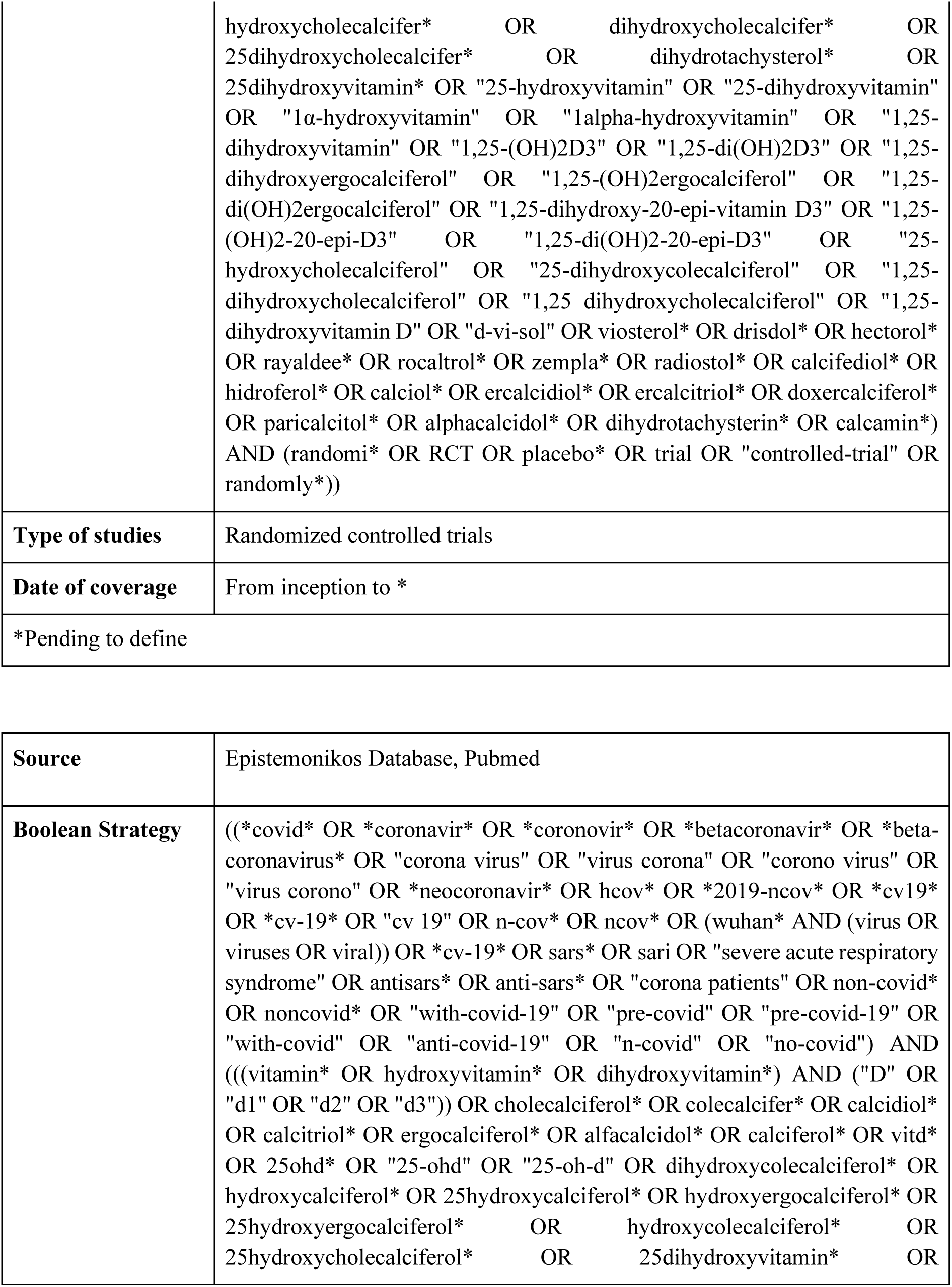
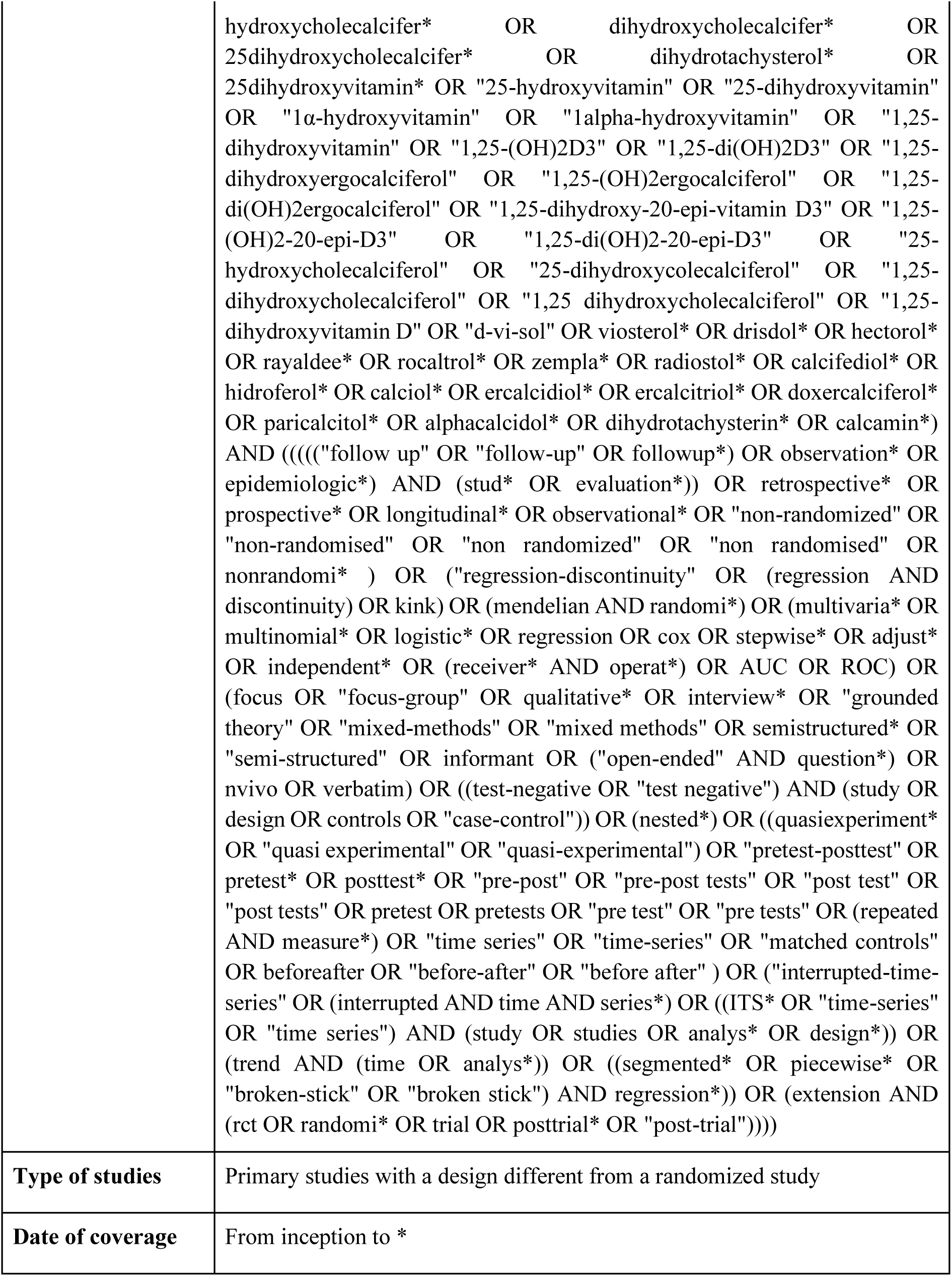
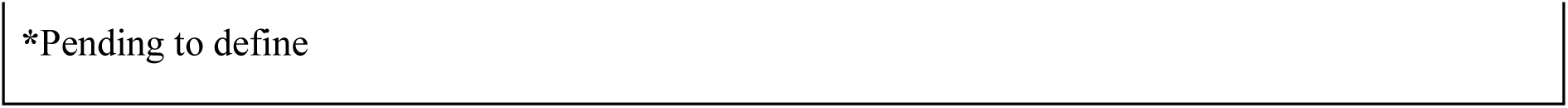
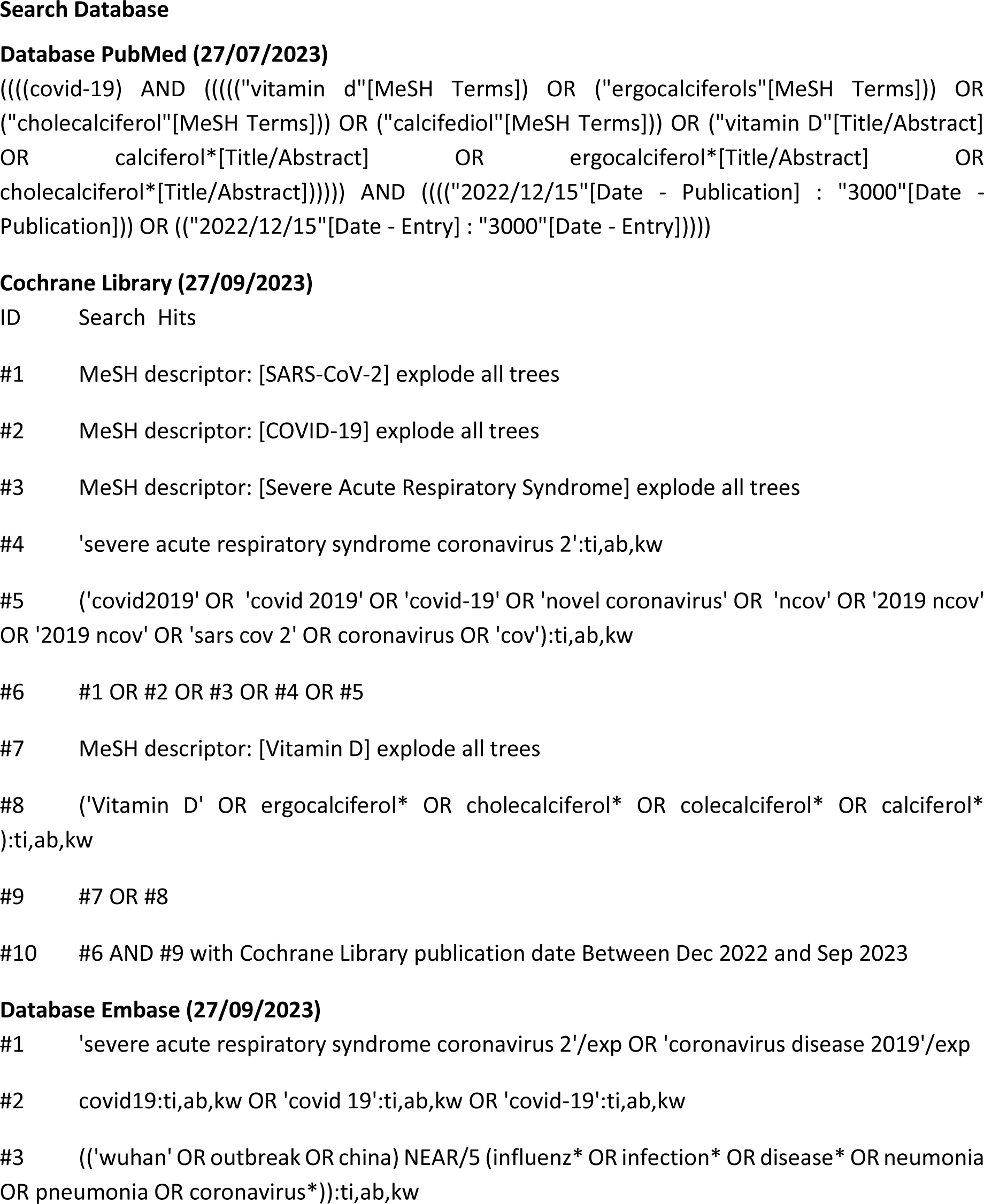
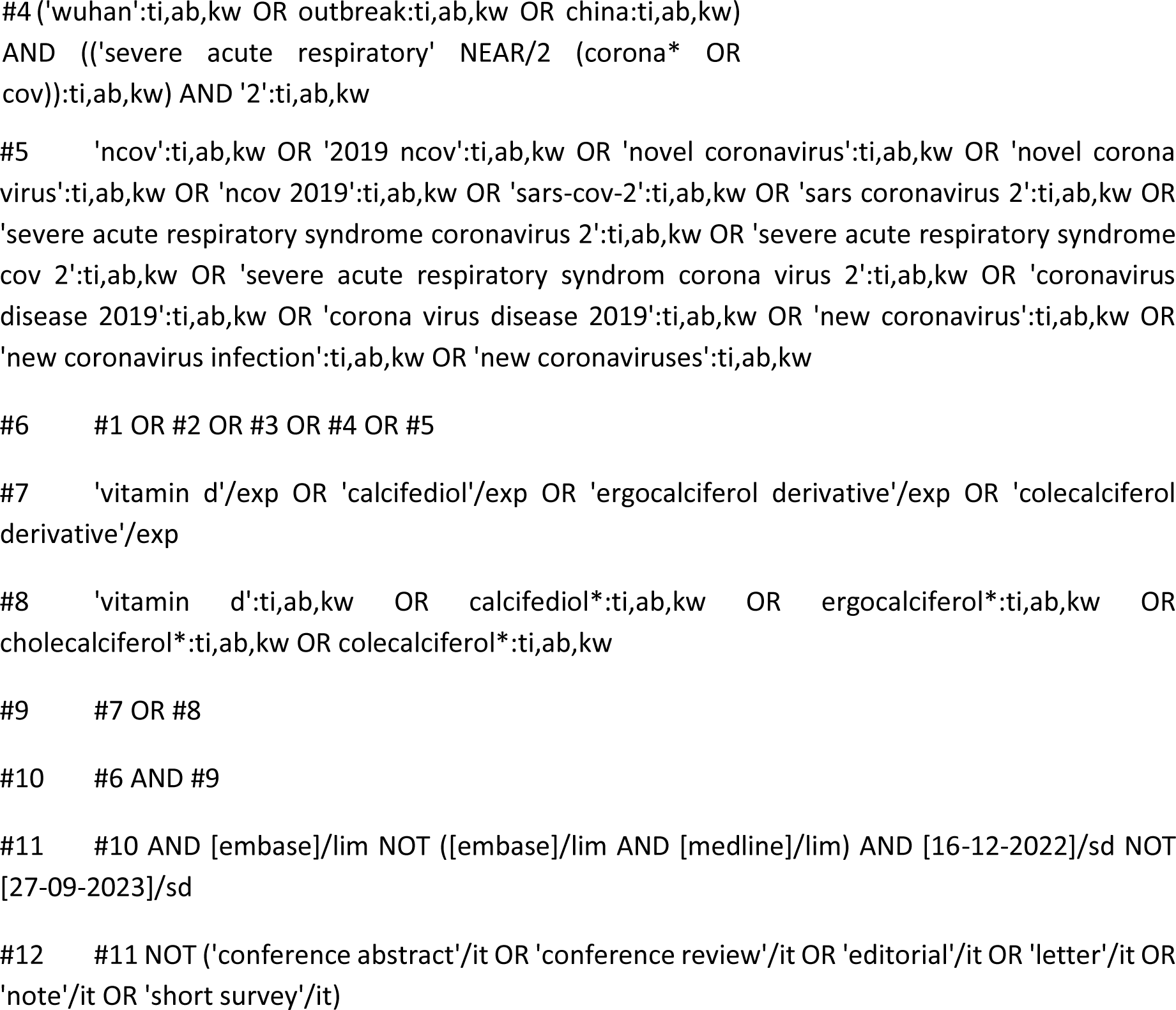

## Appendix 2 Characteristics of included studies and baseline characteristics

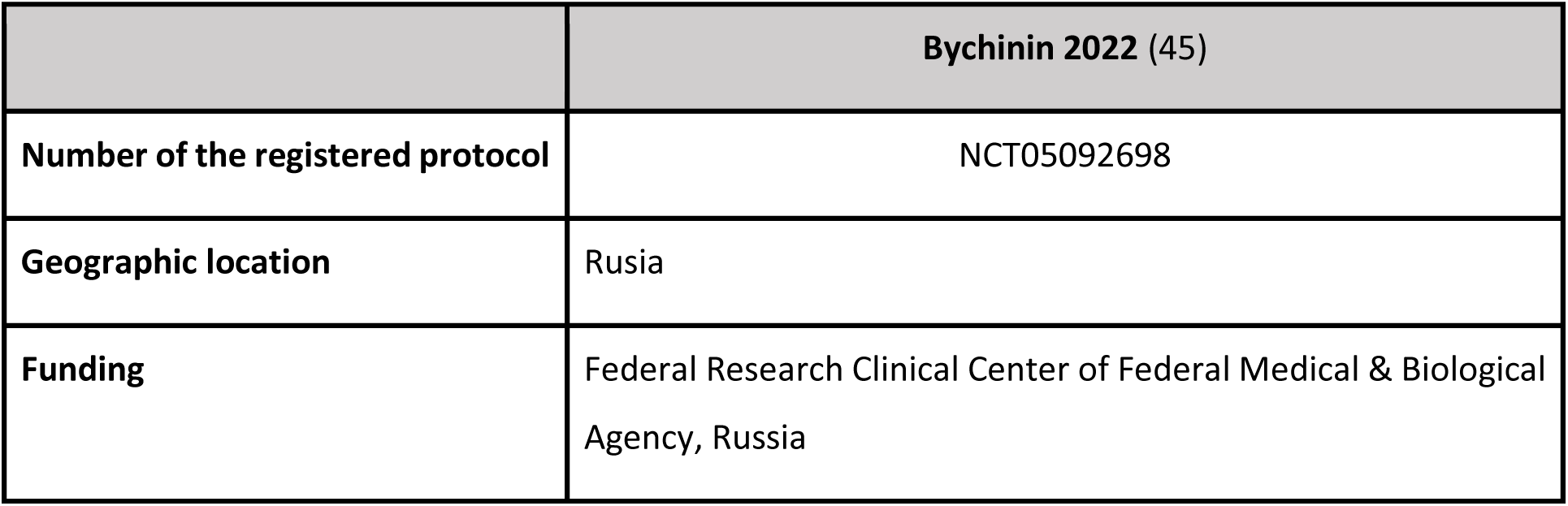
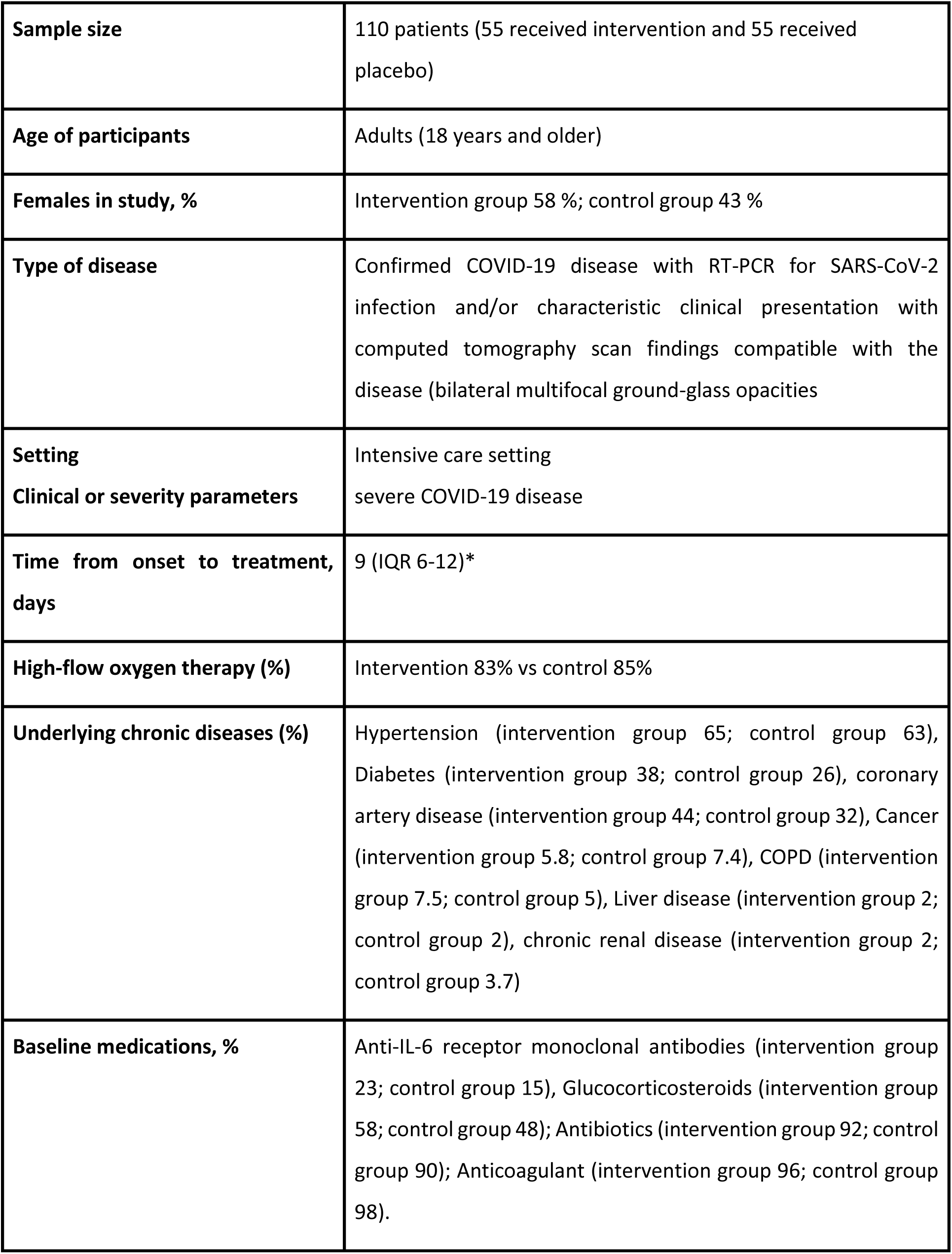
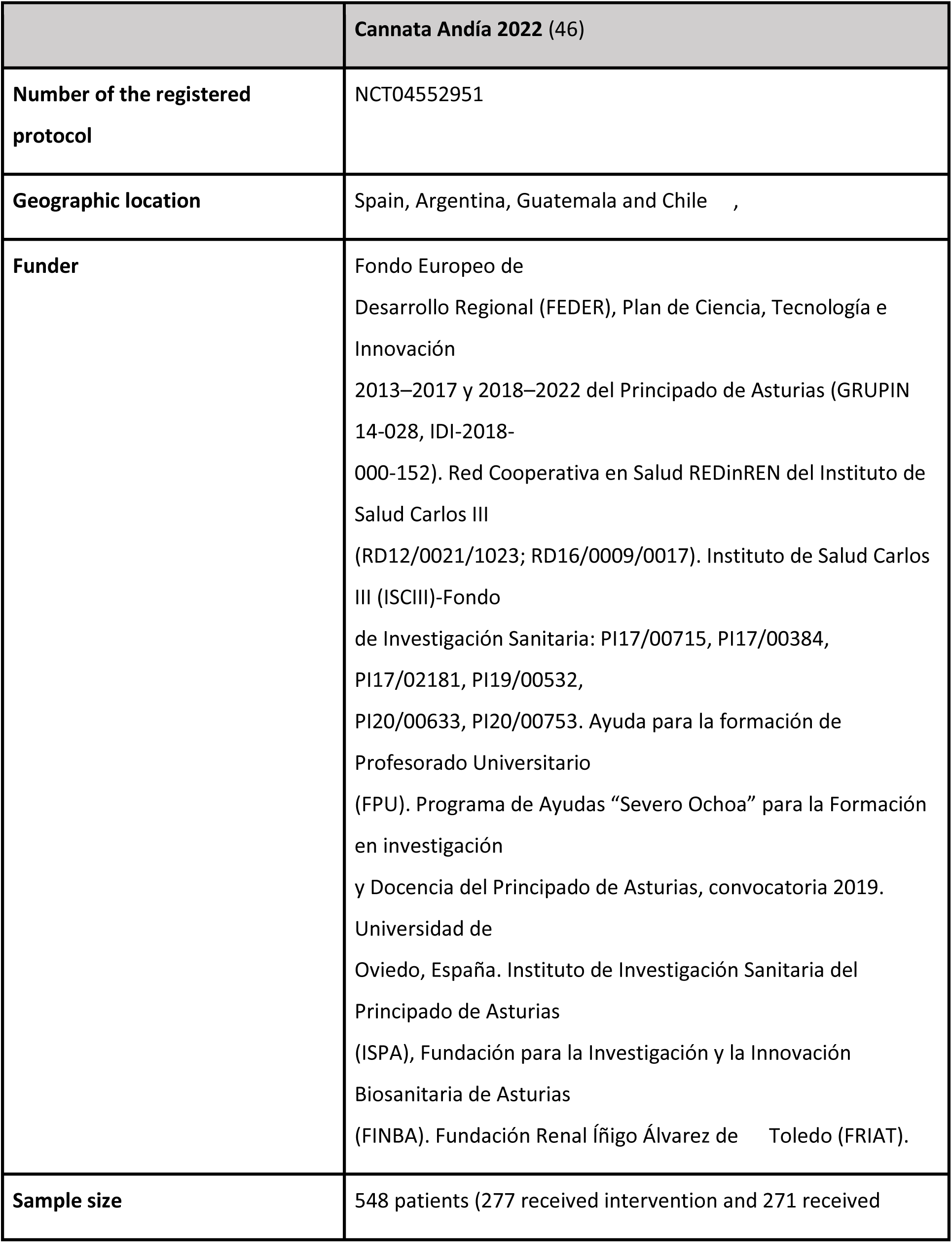
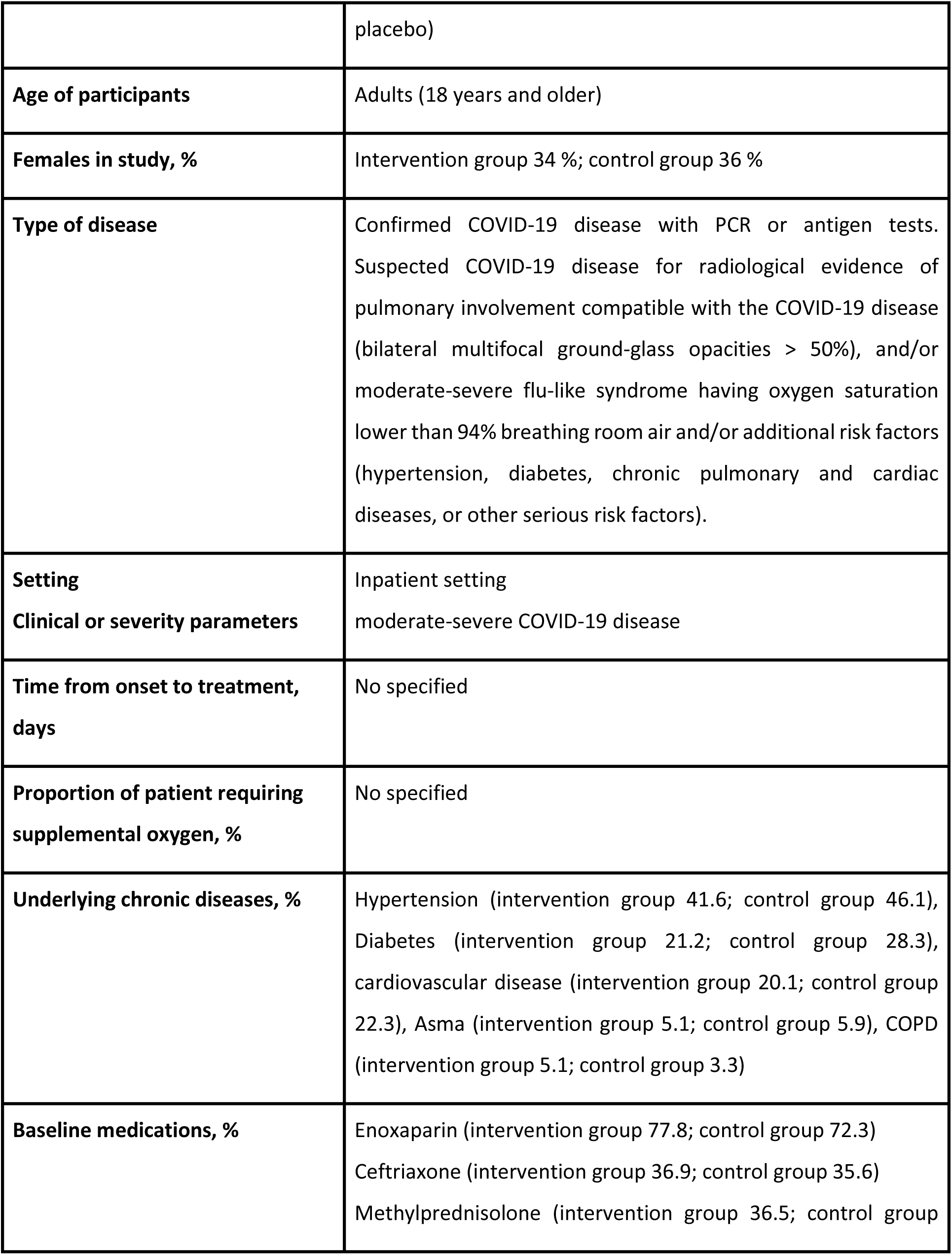
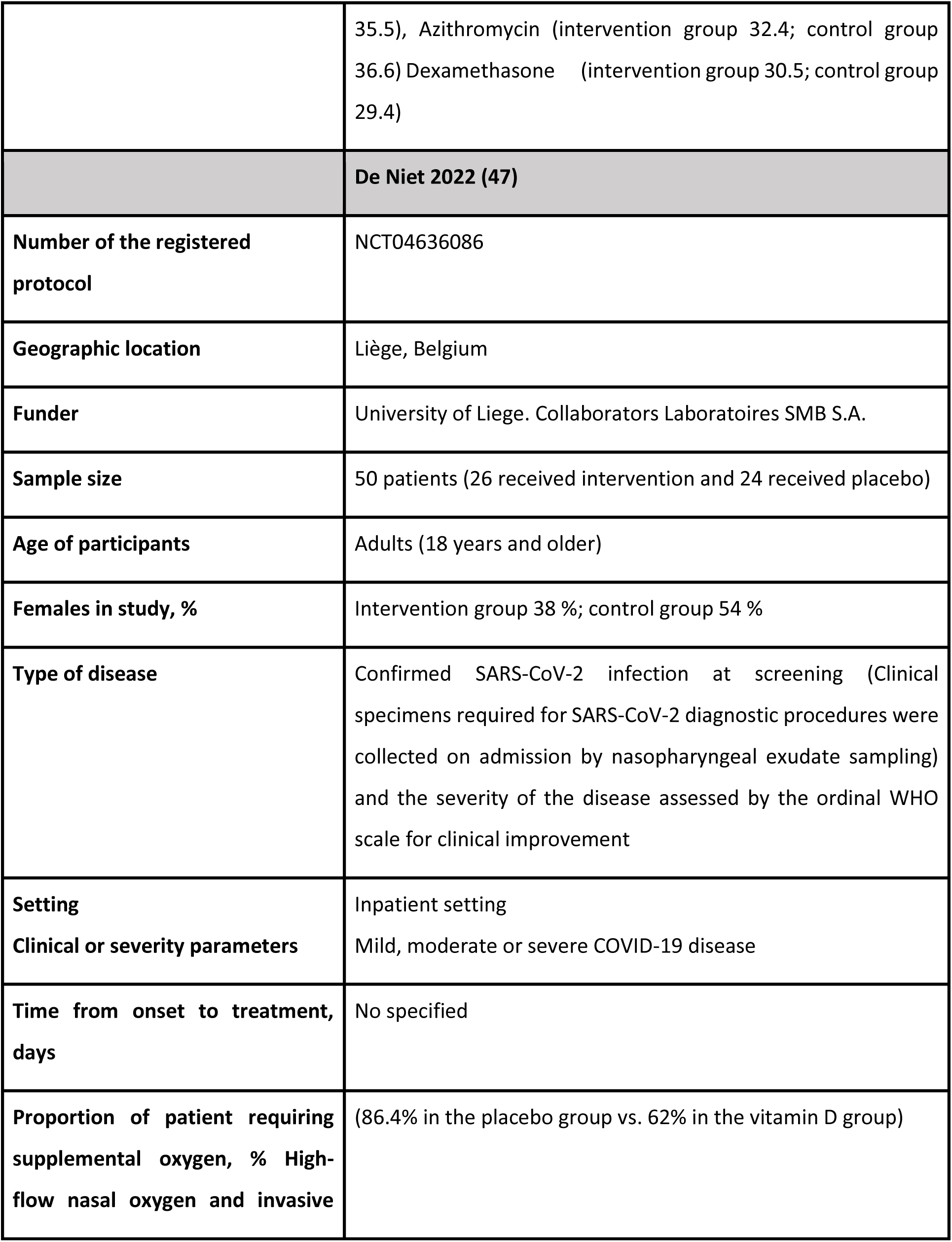
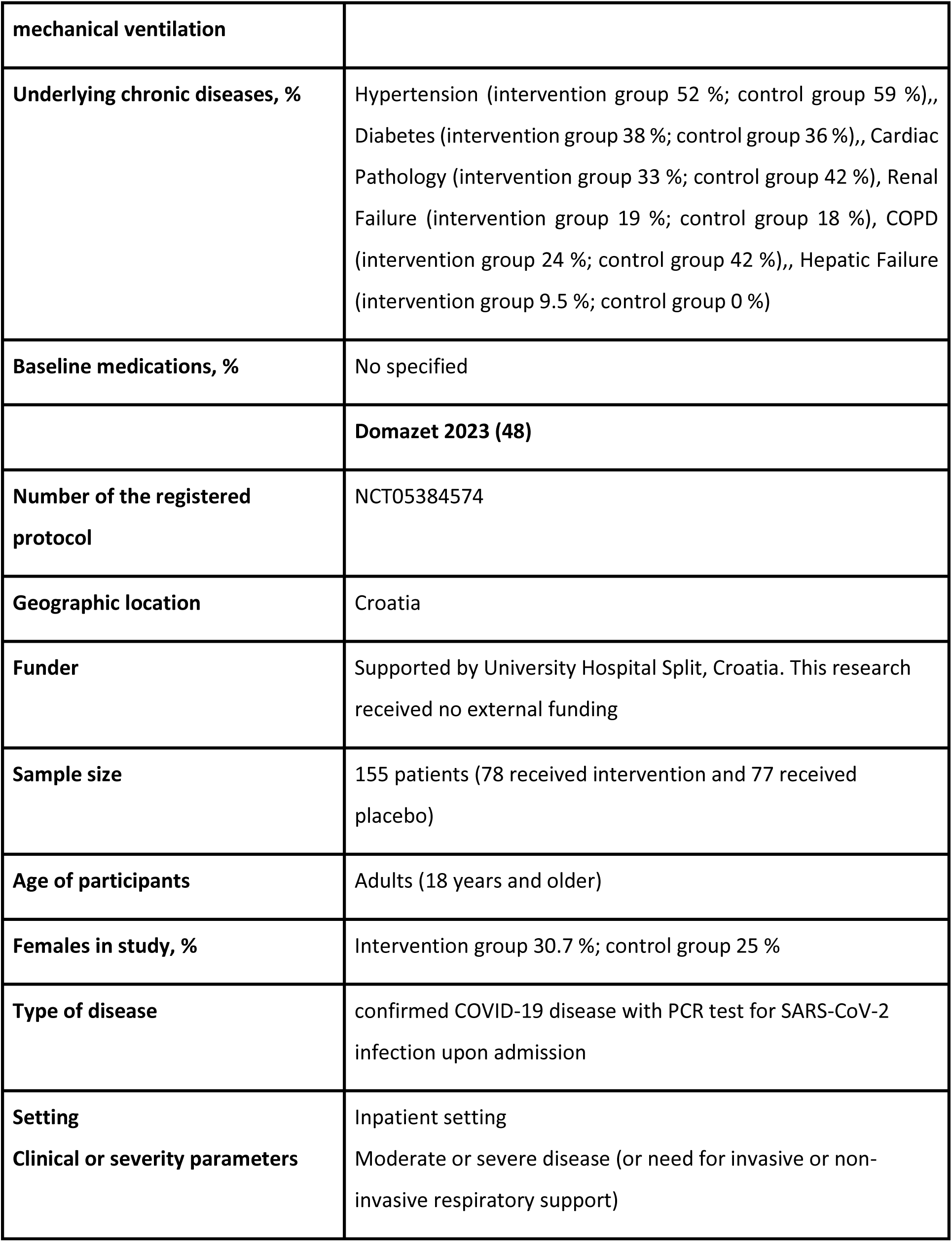
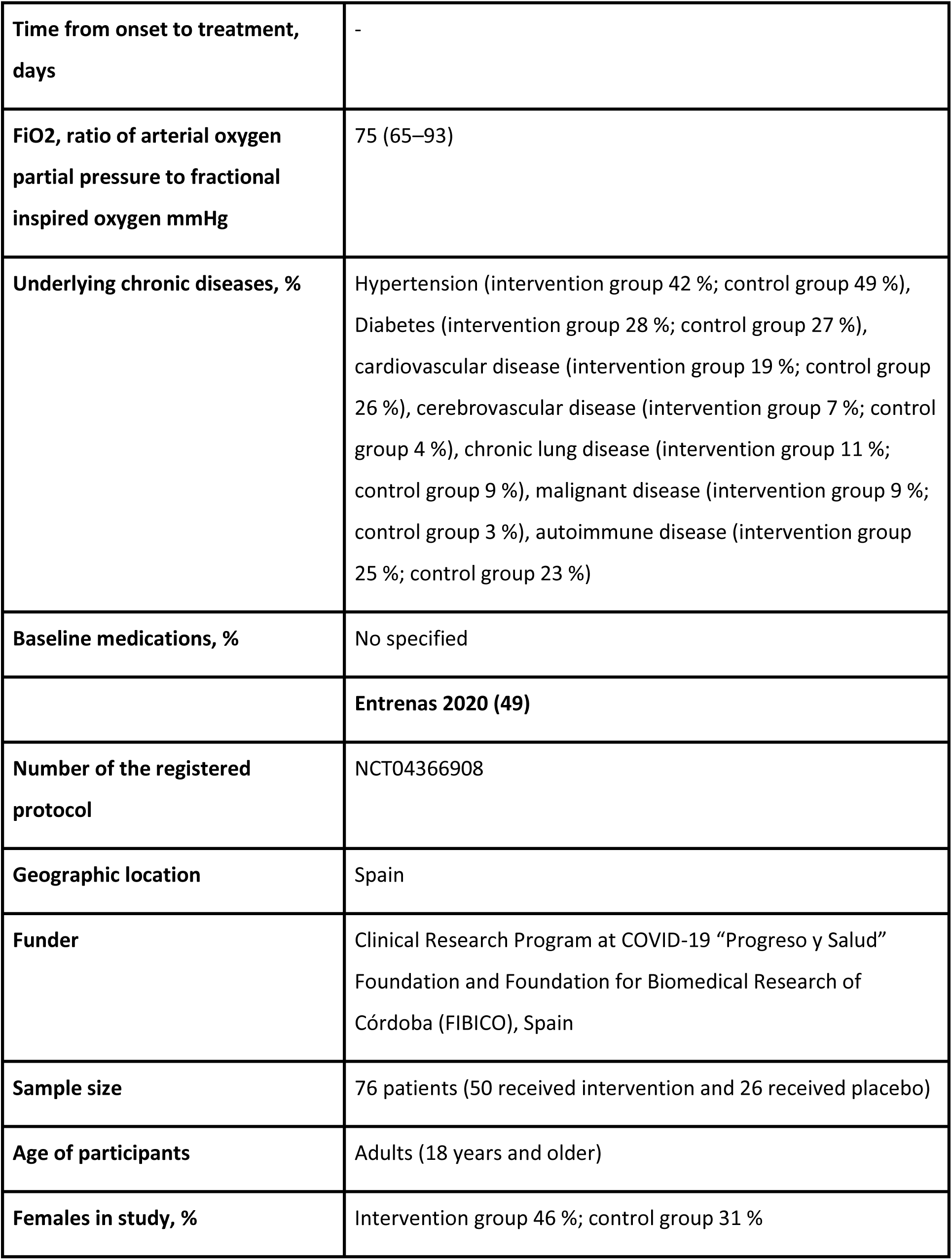
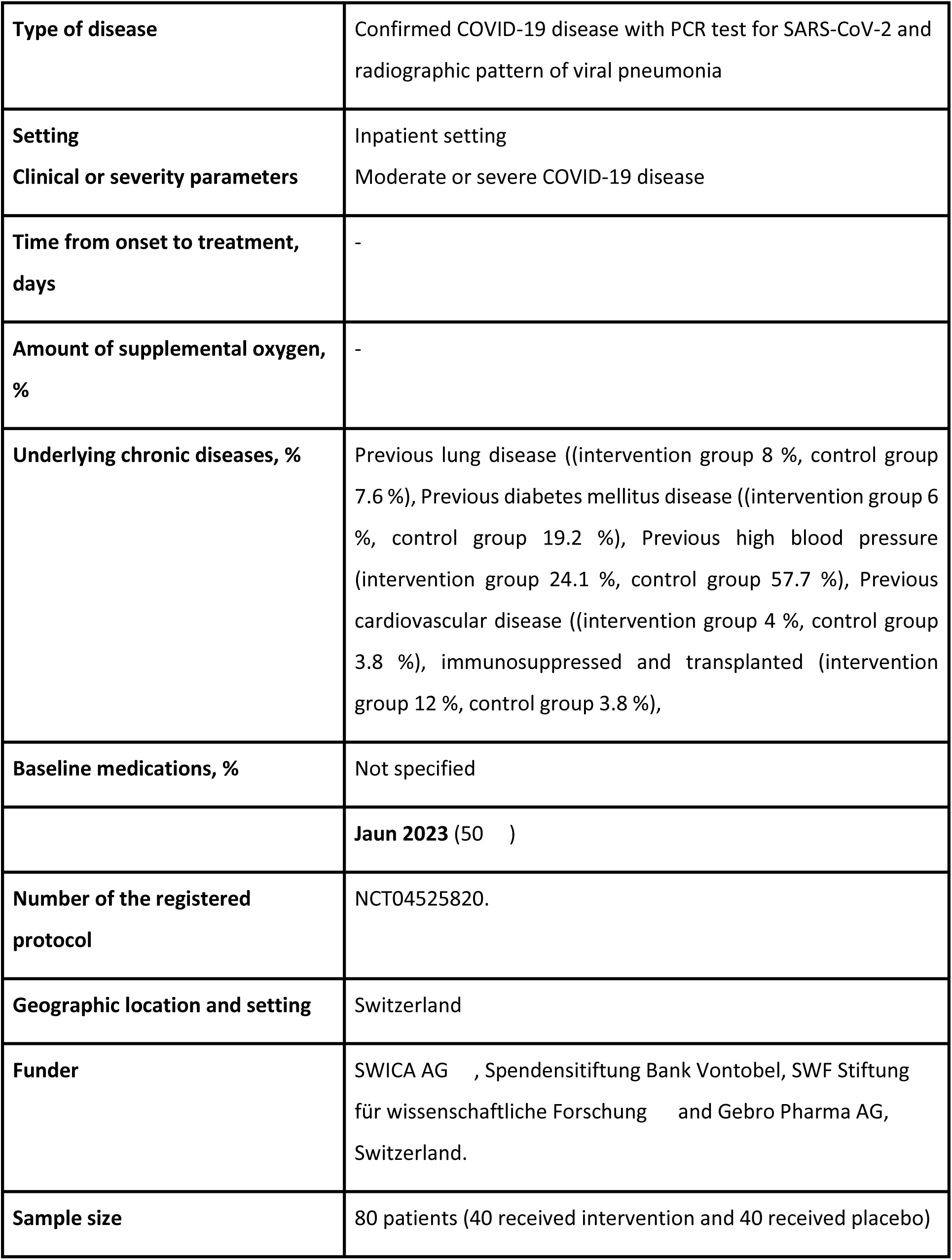
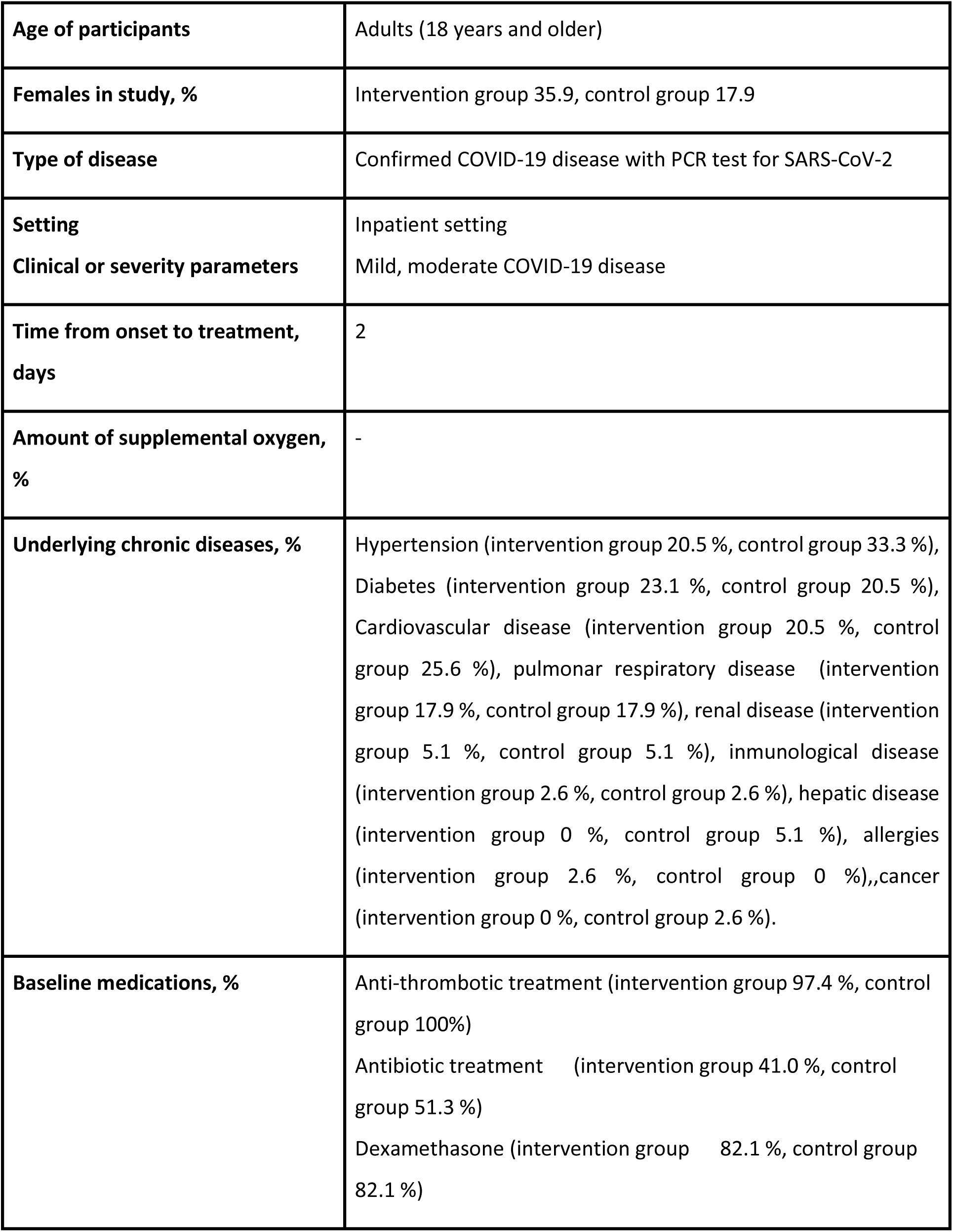
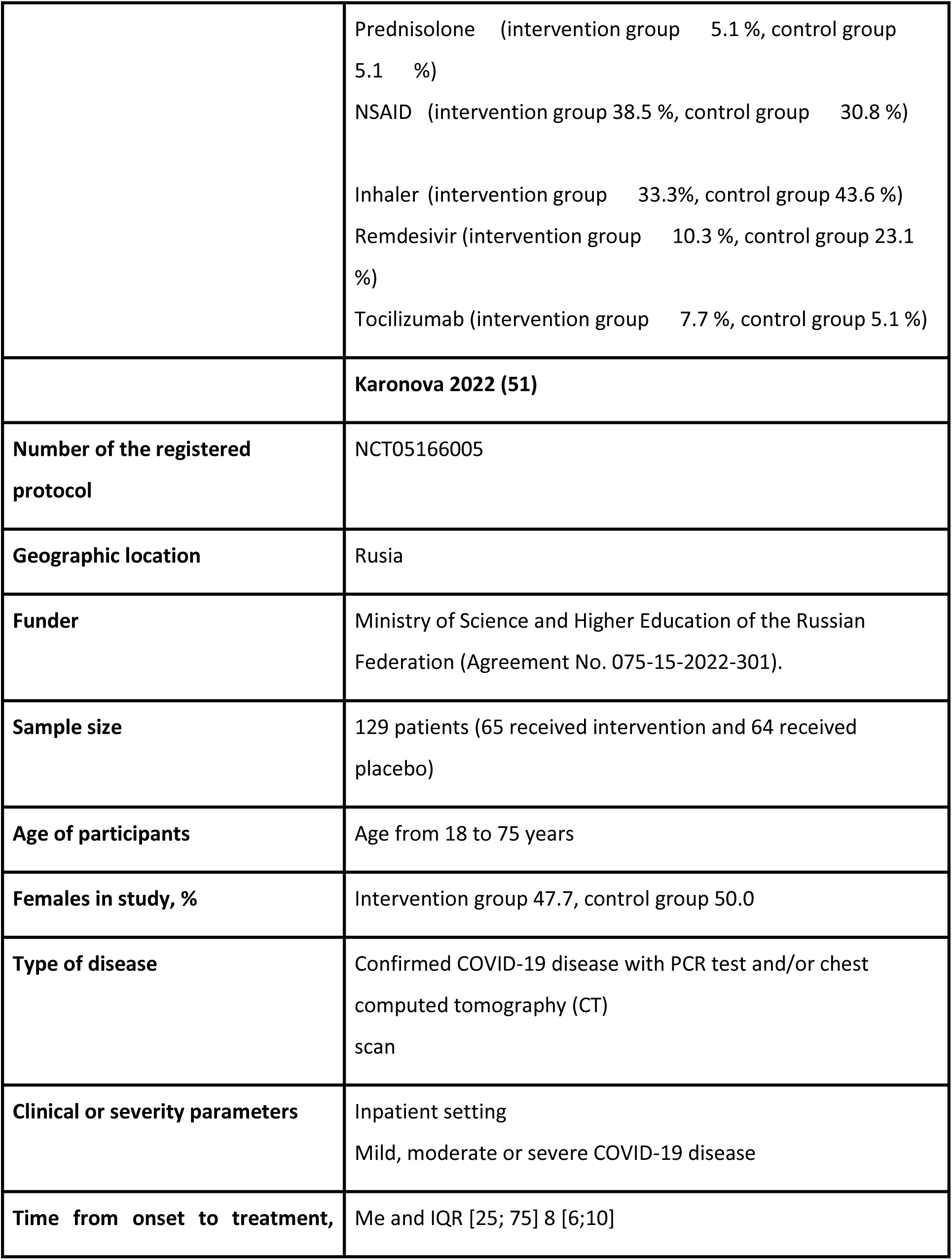
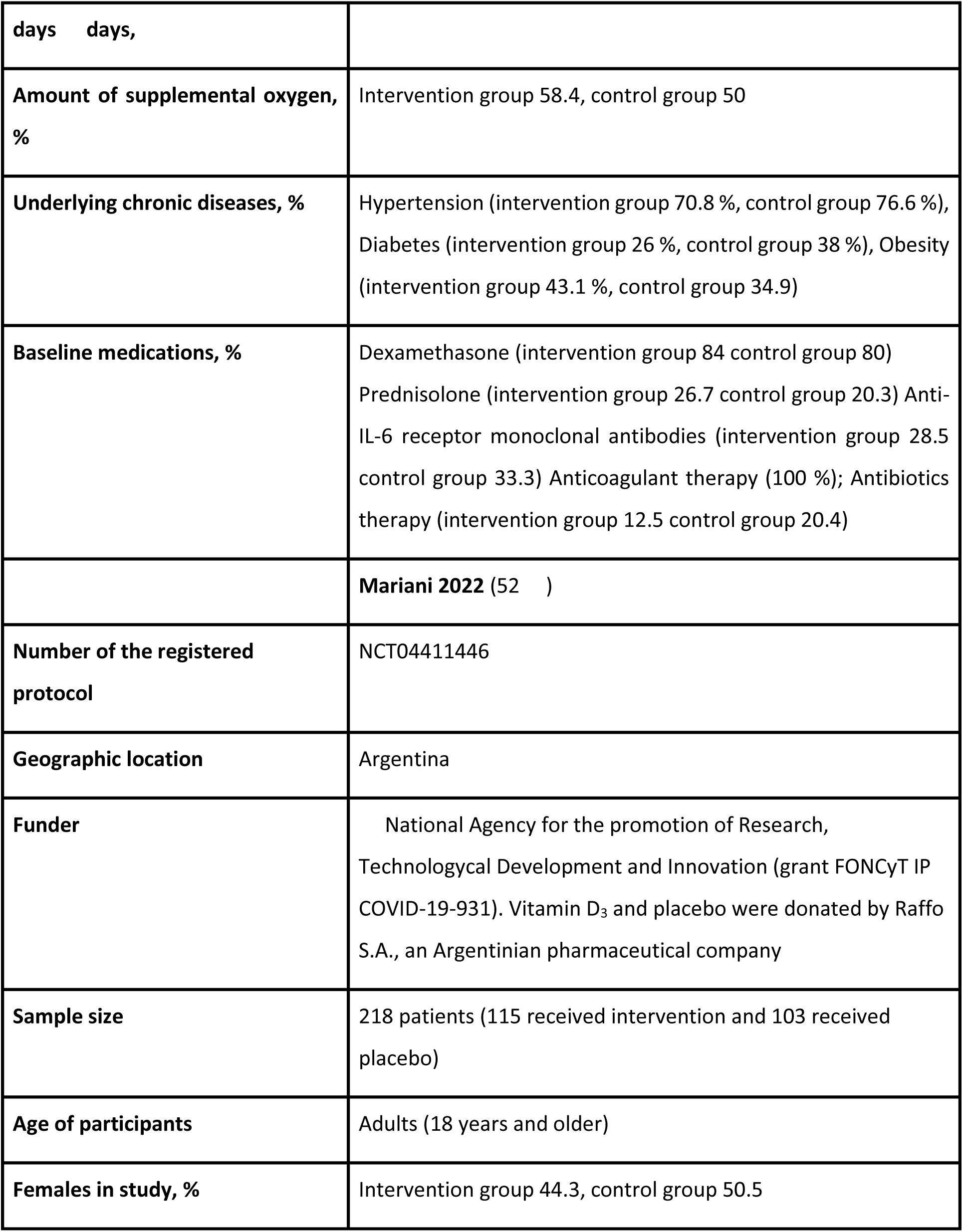
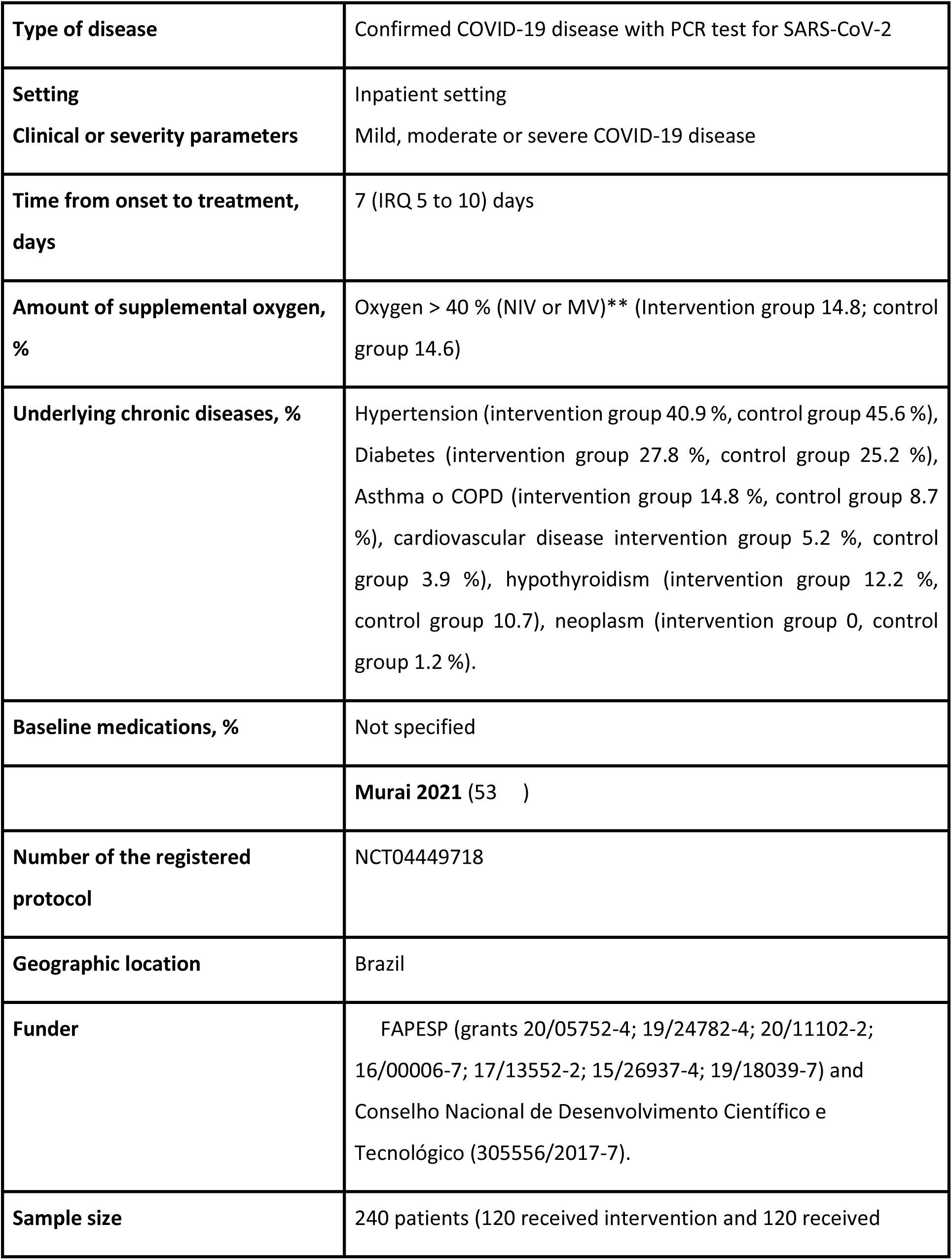
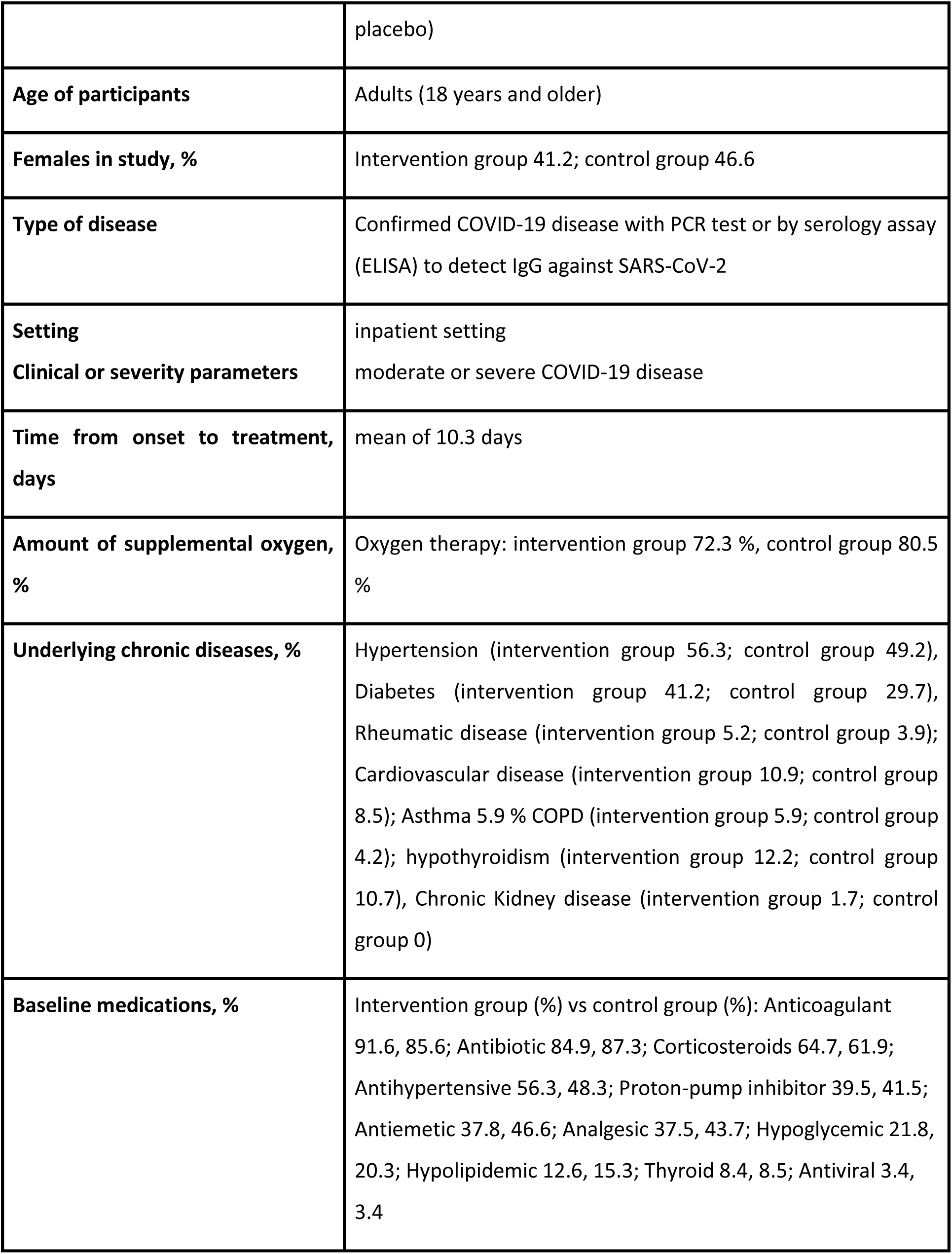
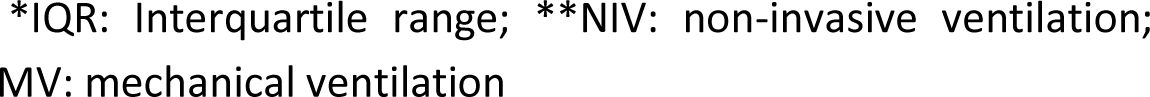

## Appendix 3. List of included, excluded and ongoing studies

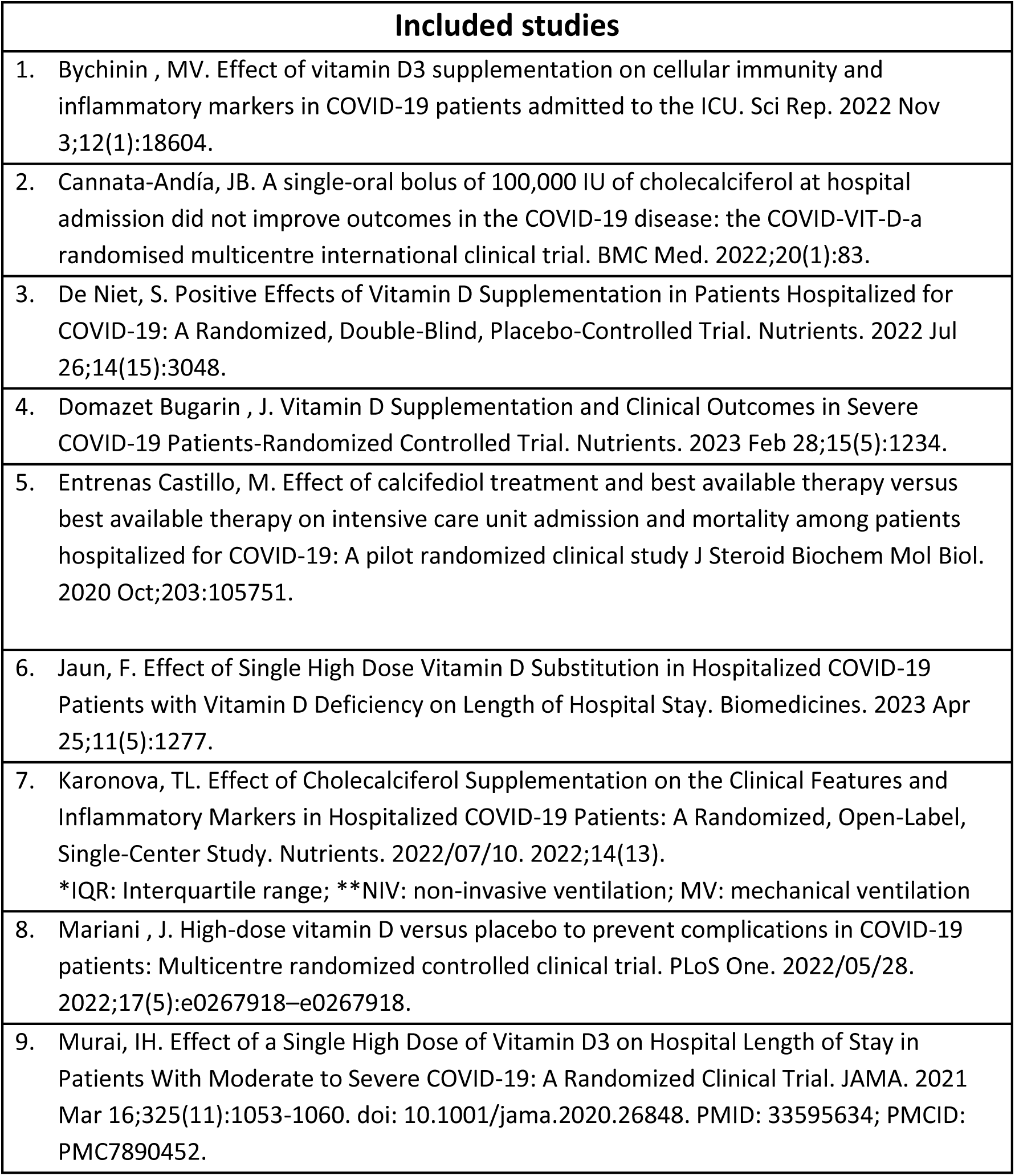
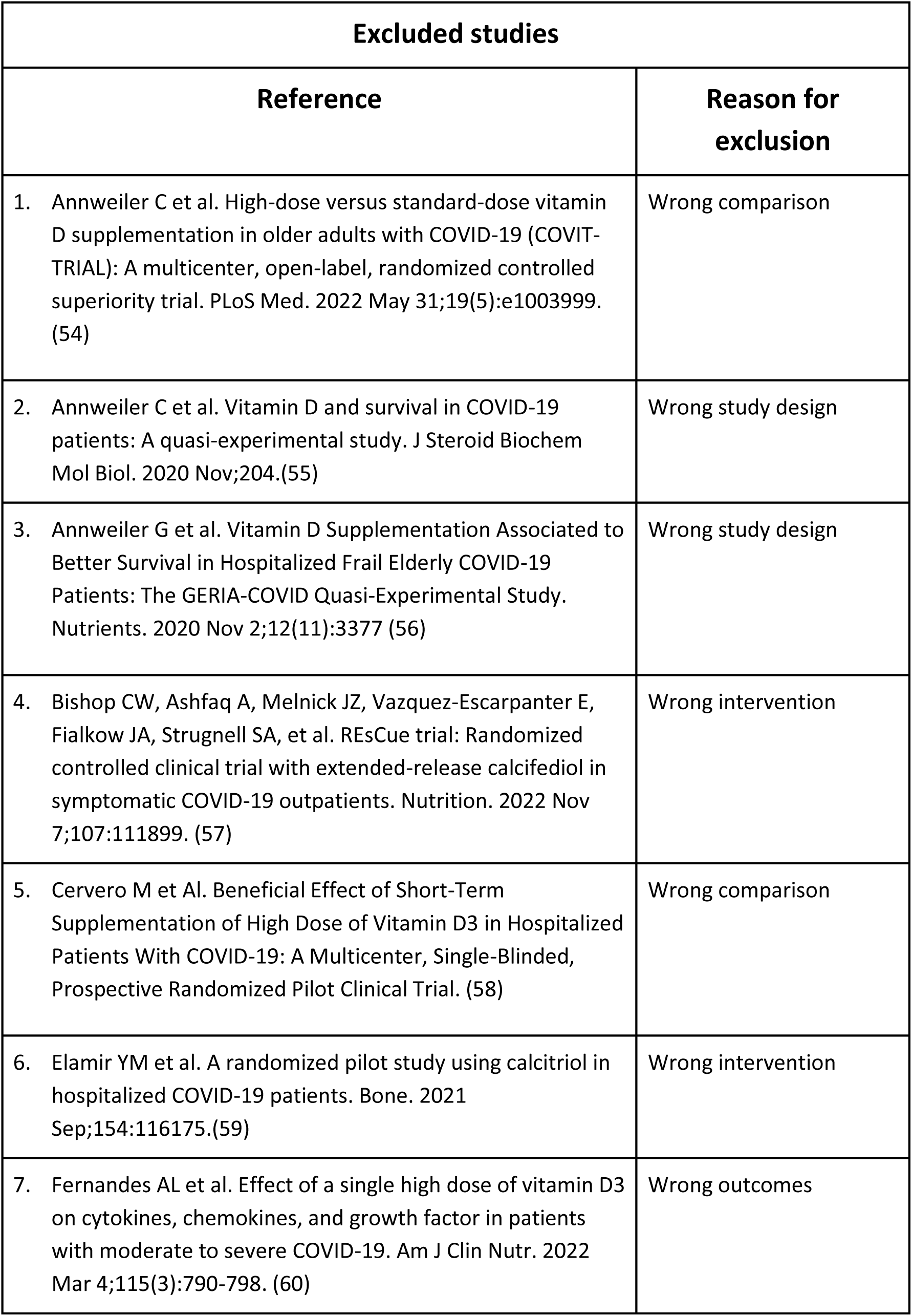
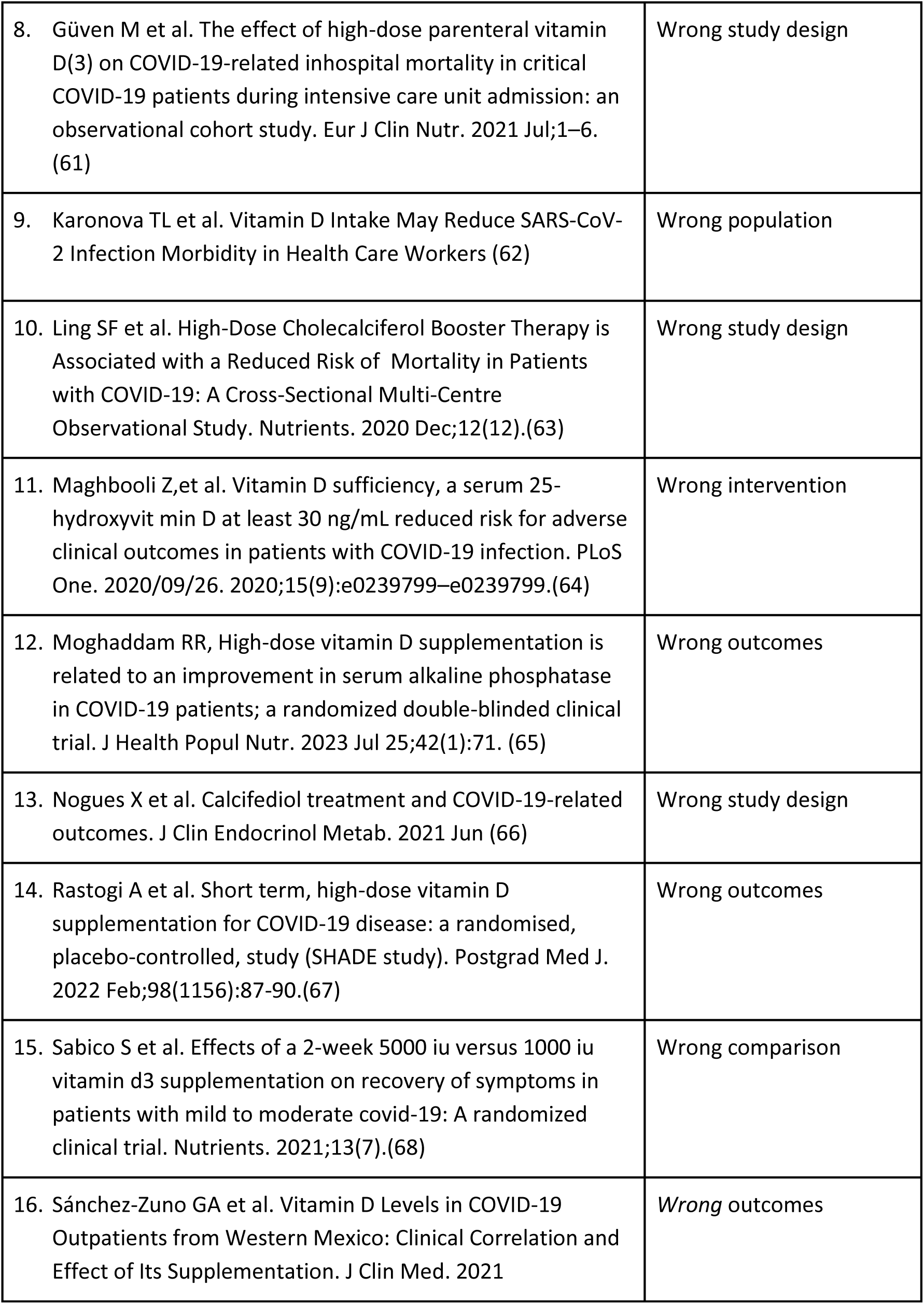
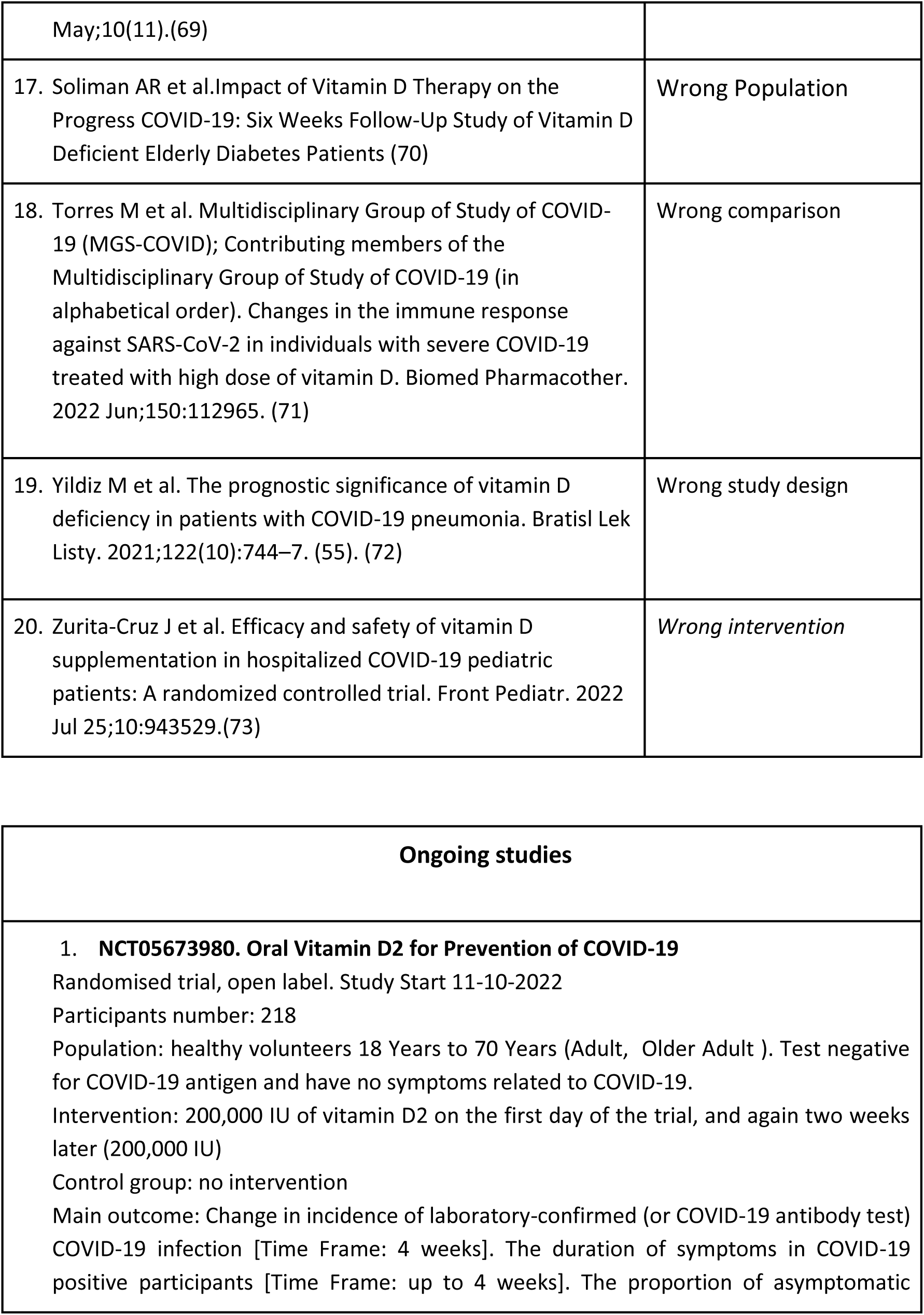
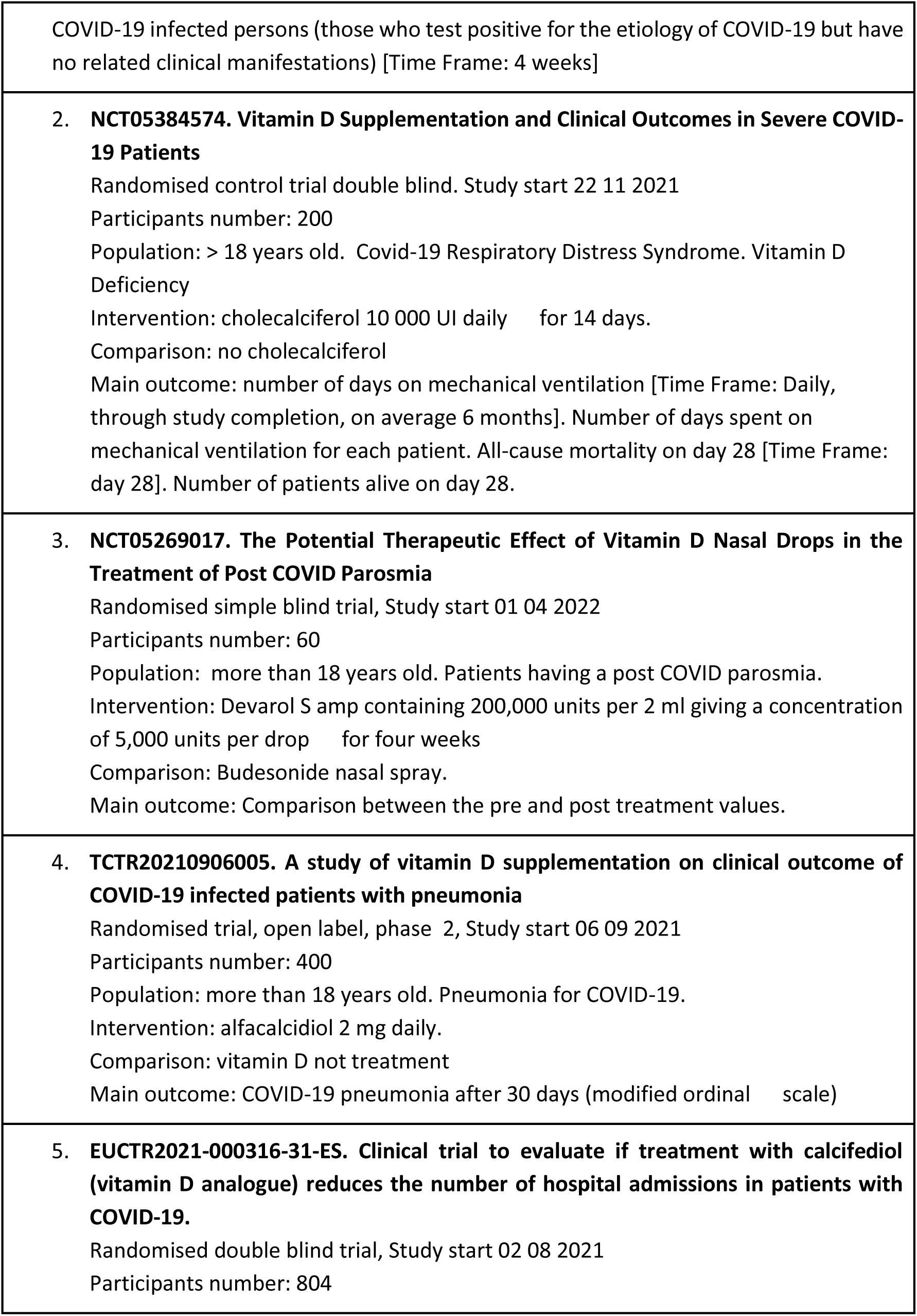
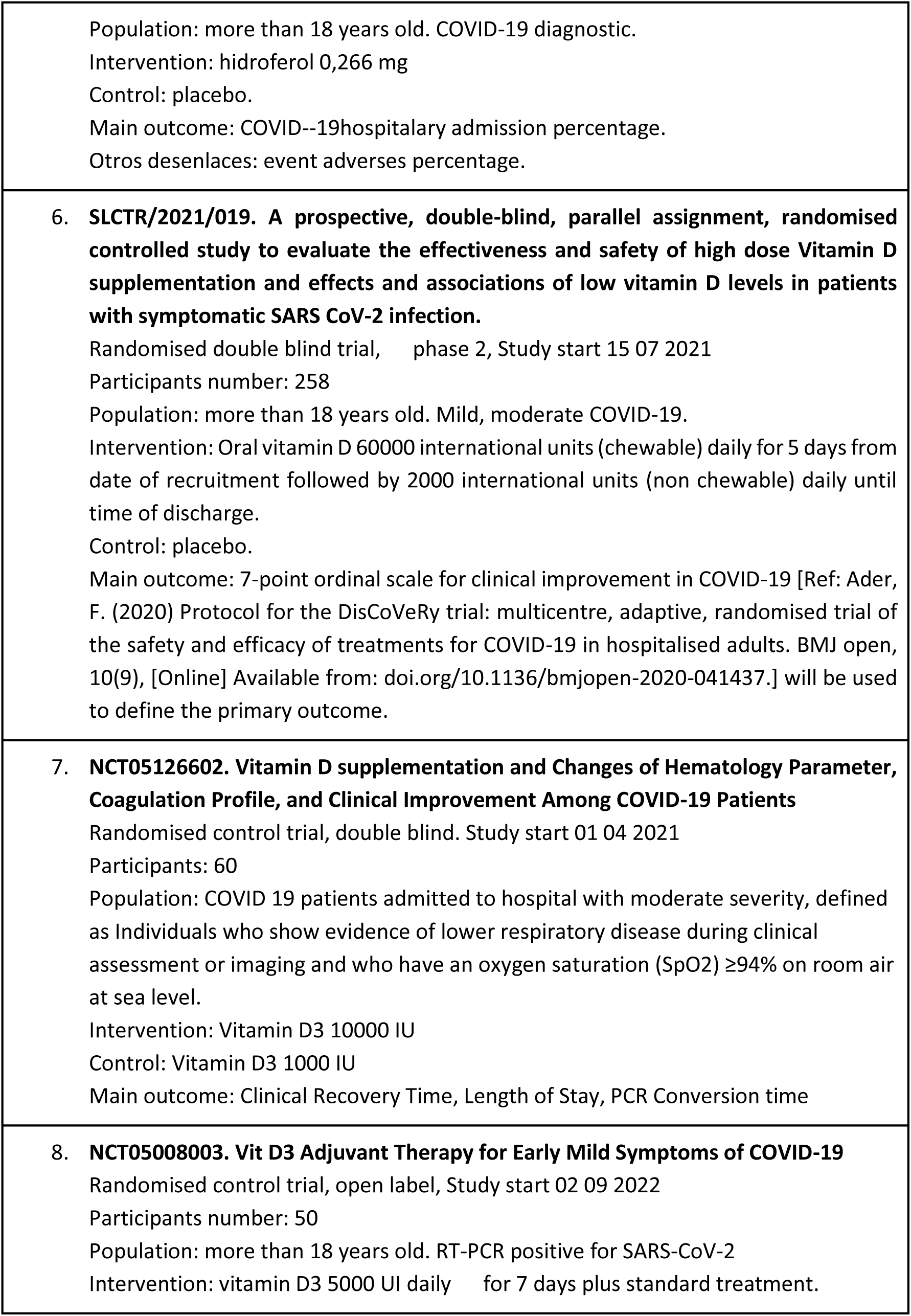
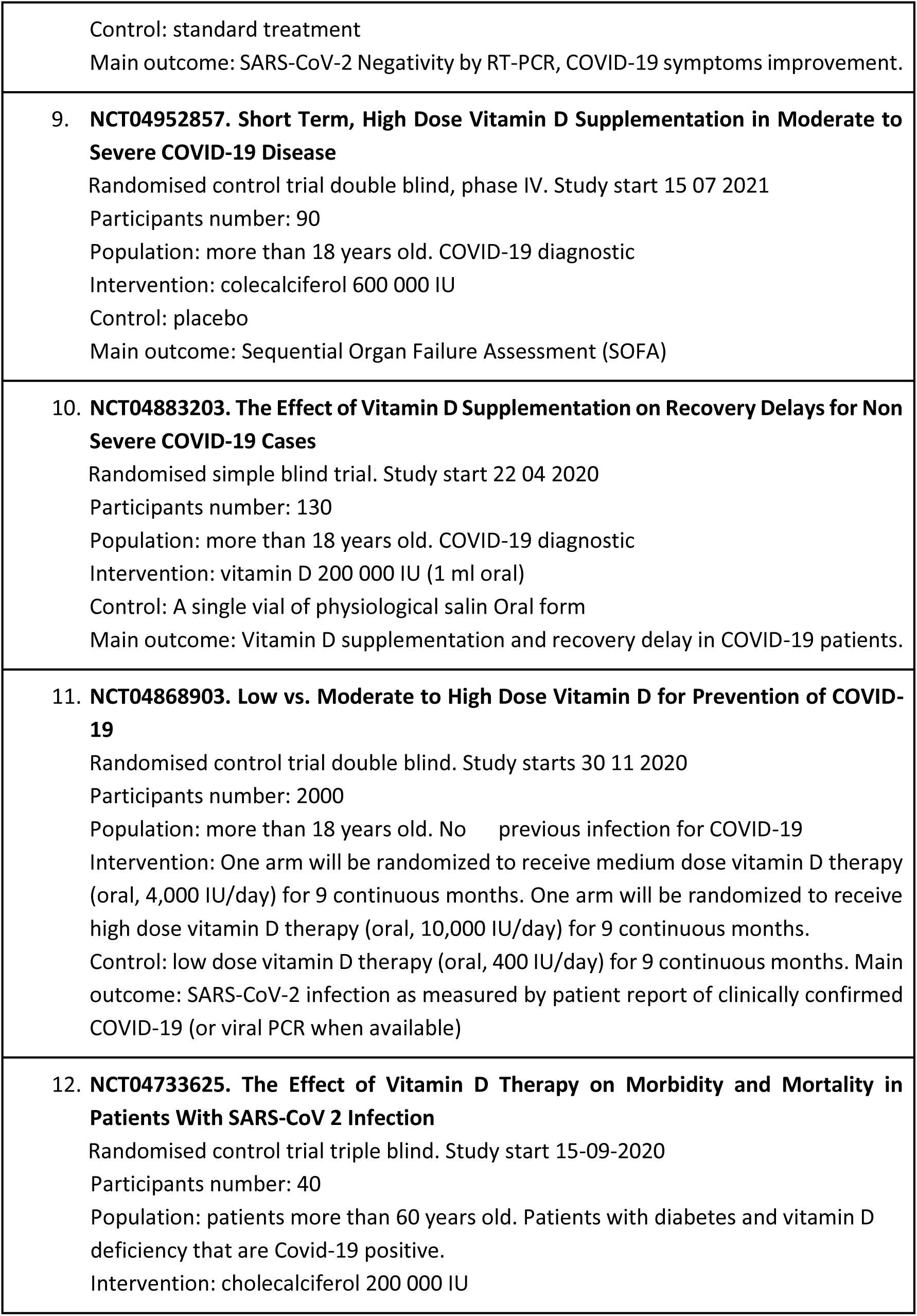
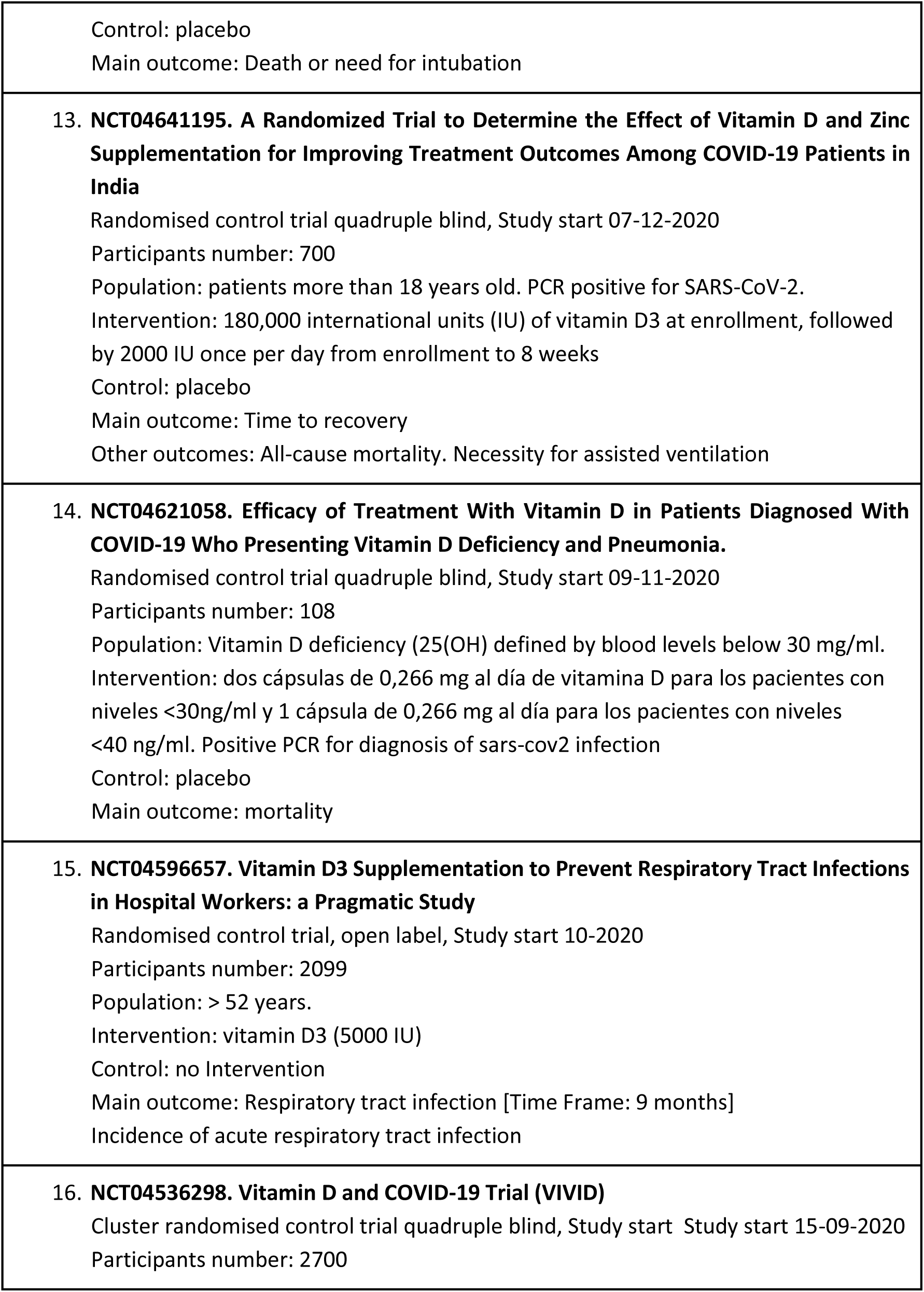
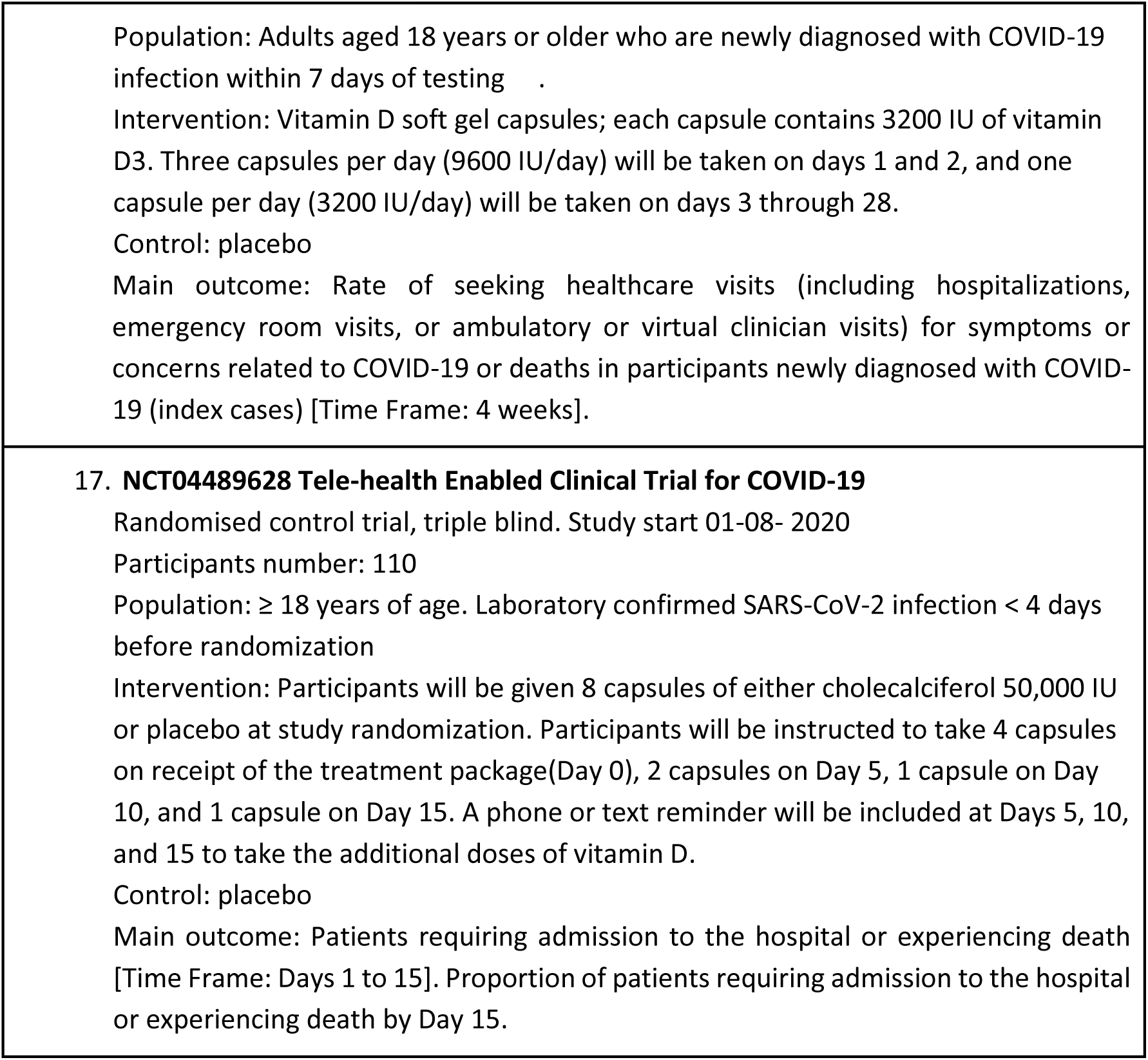
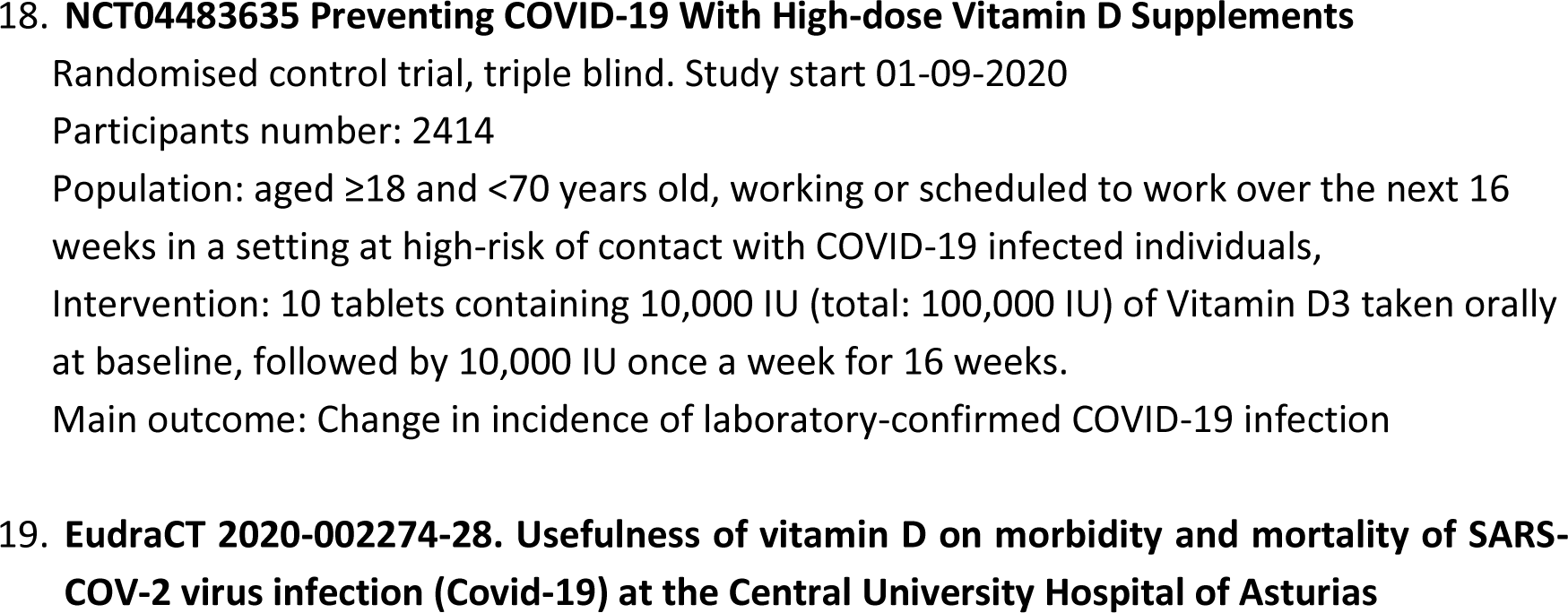
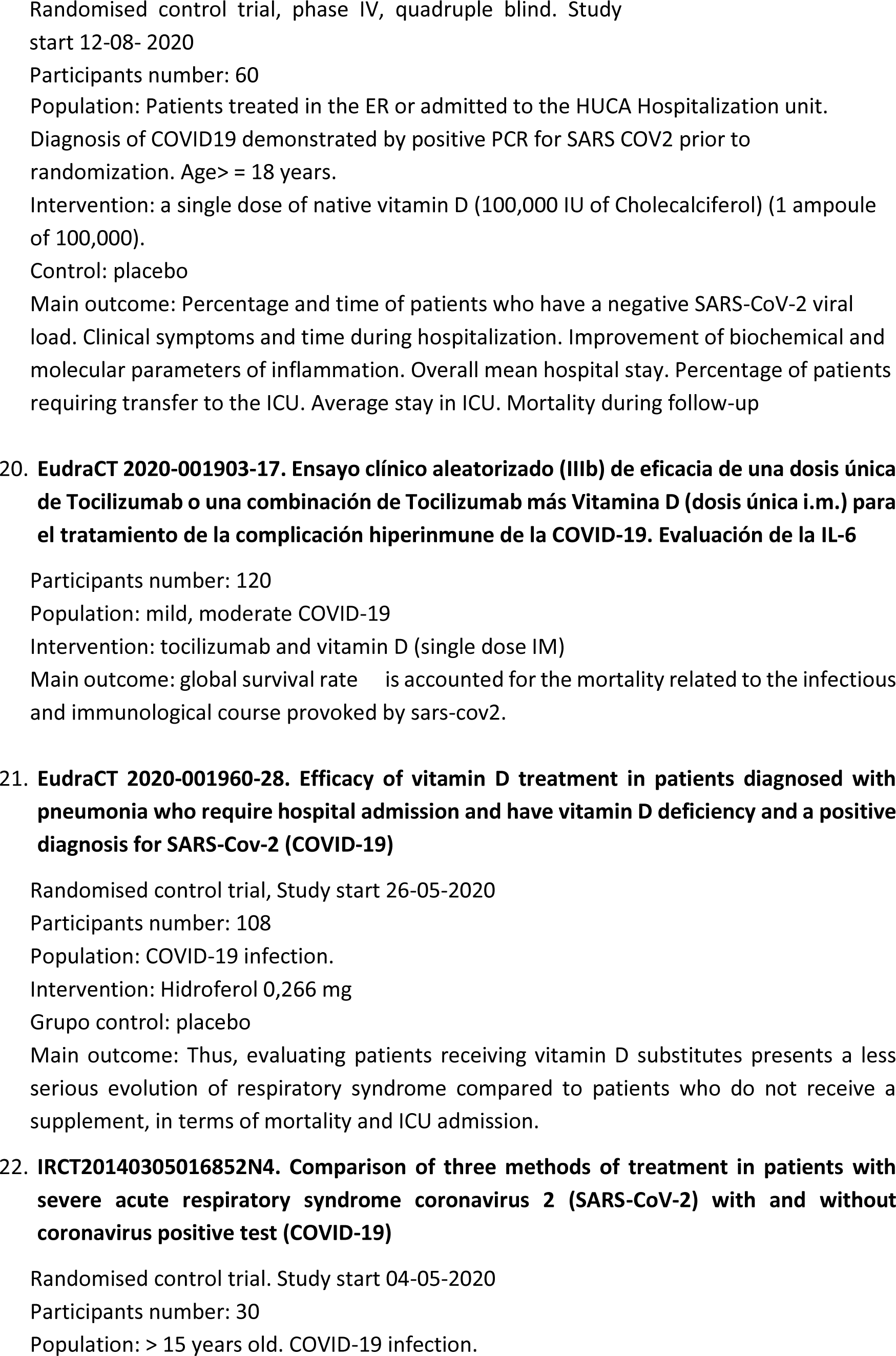
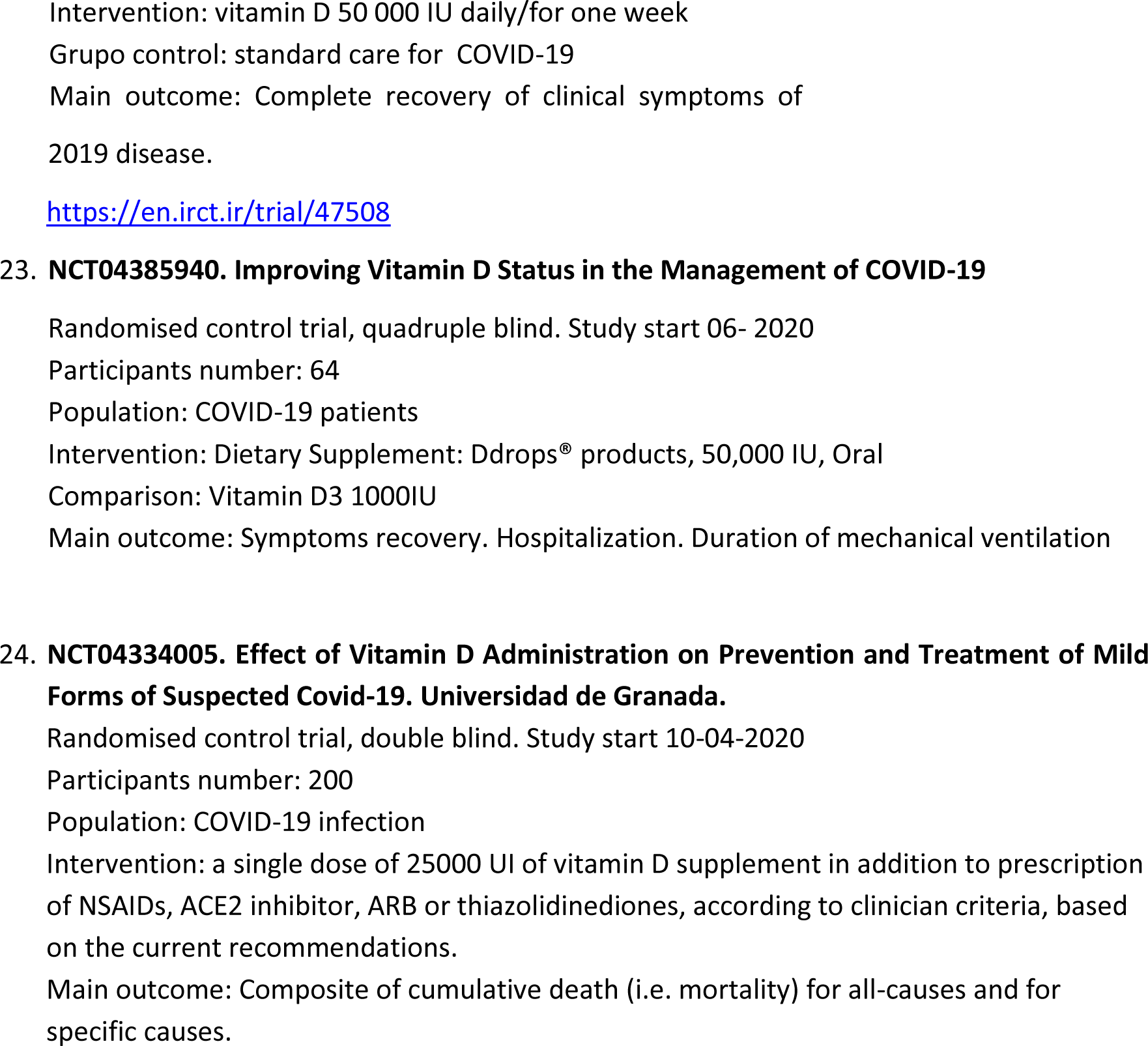

## Appendix 4. Risk of bias

**Figure 8.**
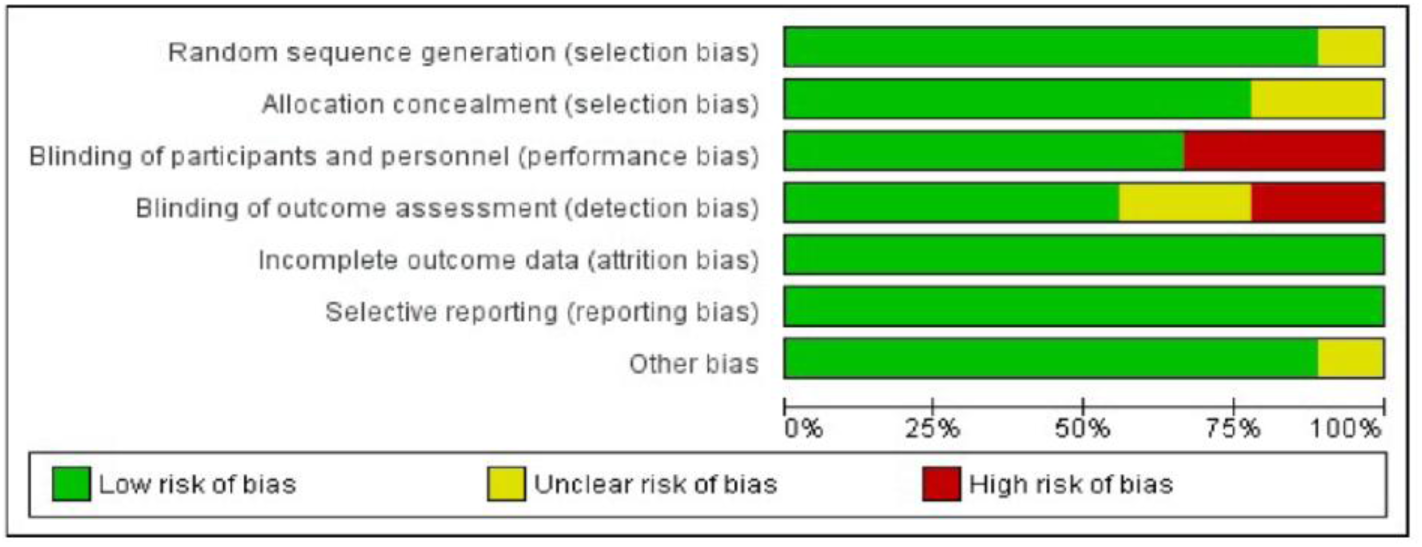
Risk of bias graph: review authors’ judgements about each risk of bias item presented as percentages across all included studies.

**Figure 9.**
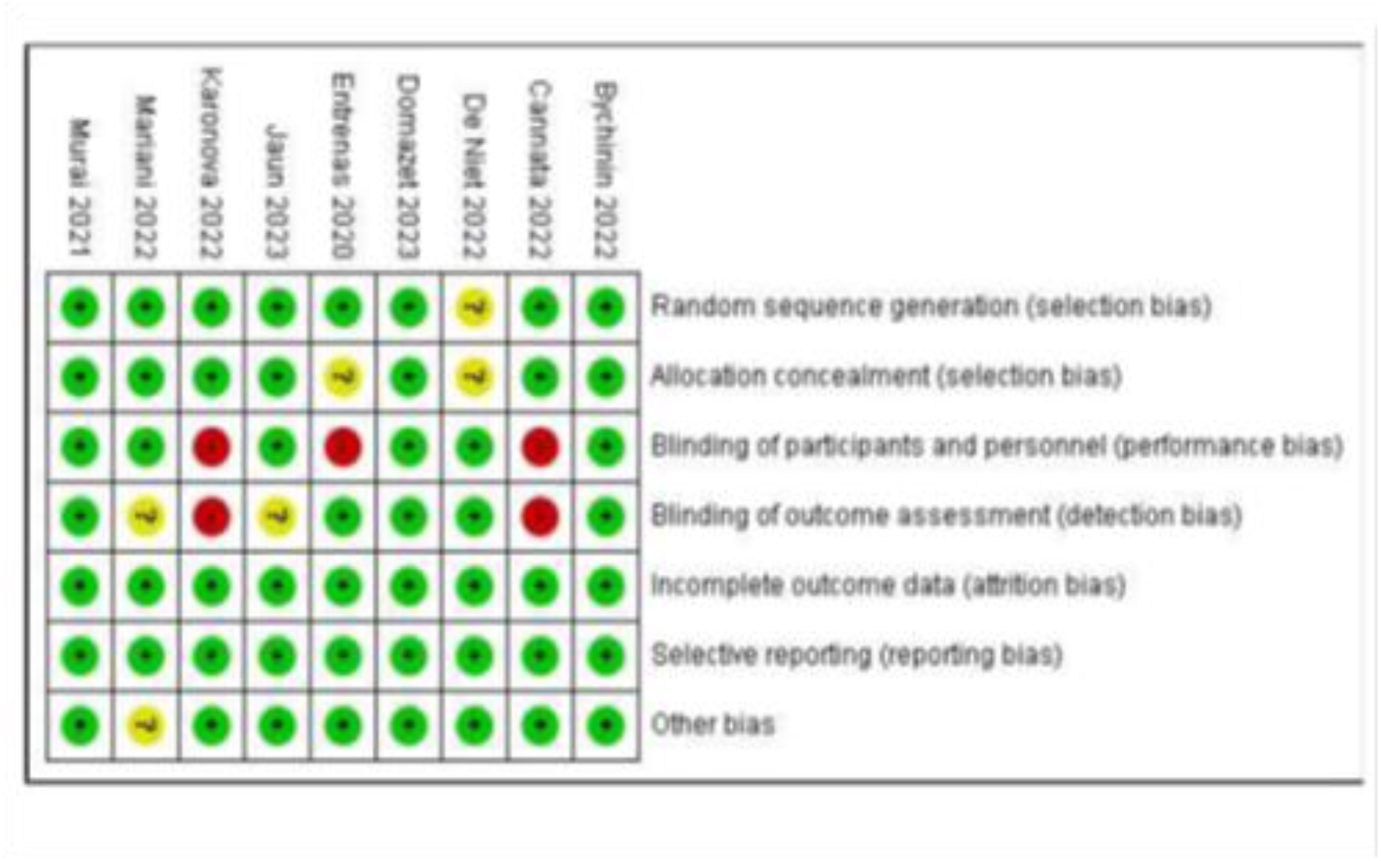
Risk of bias summary: review authors’ judgements about each risk of bias item for each included study.

## Appendix 5. Evidence profile

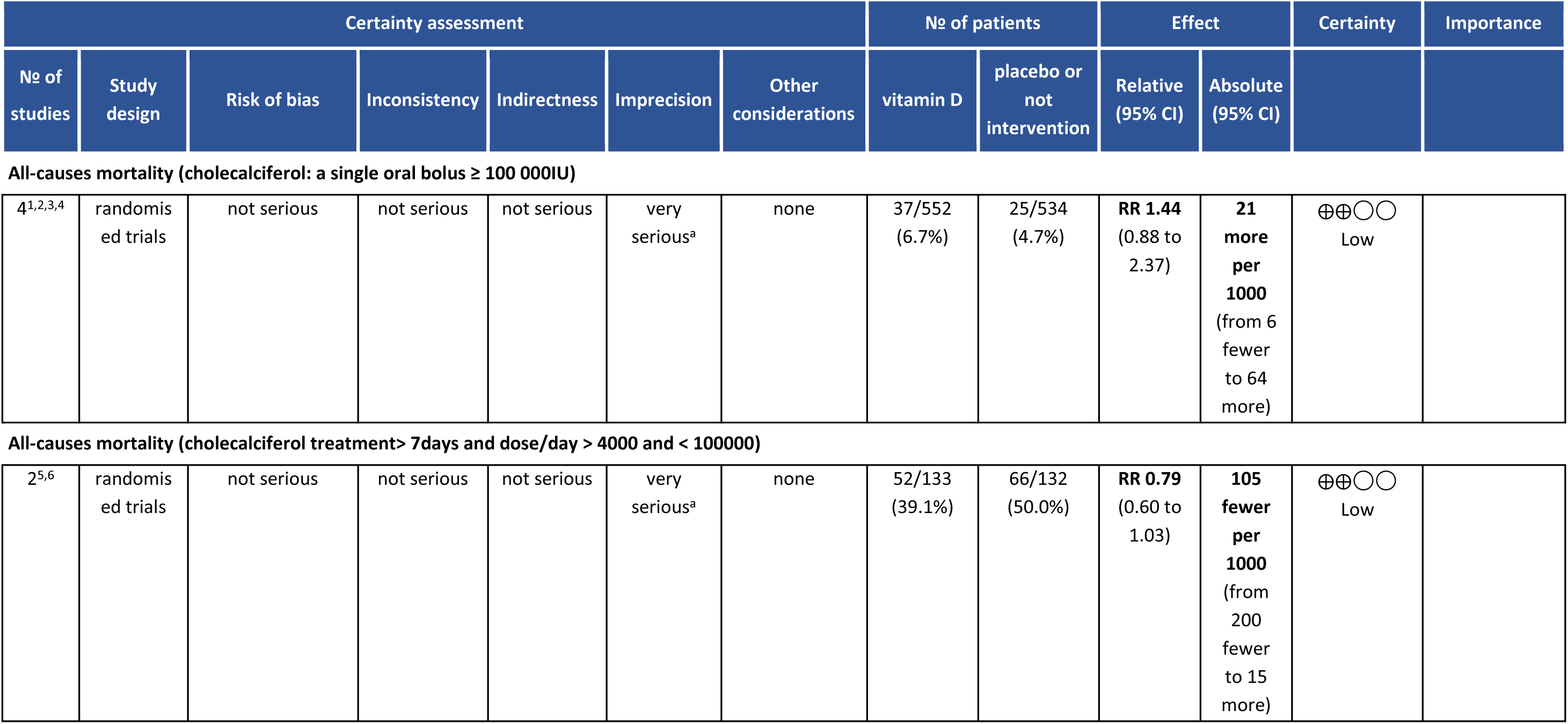
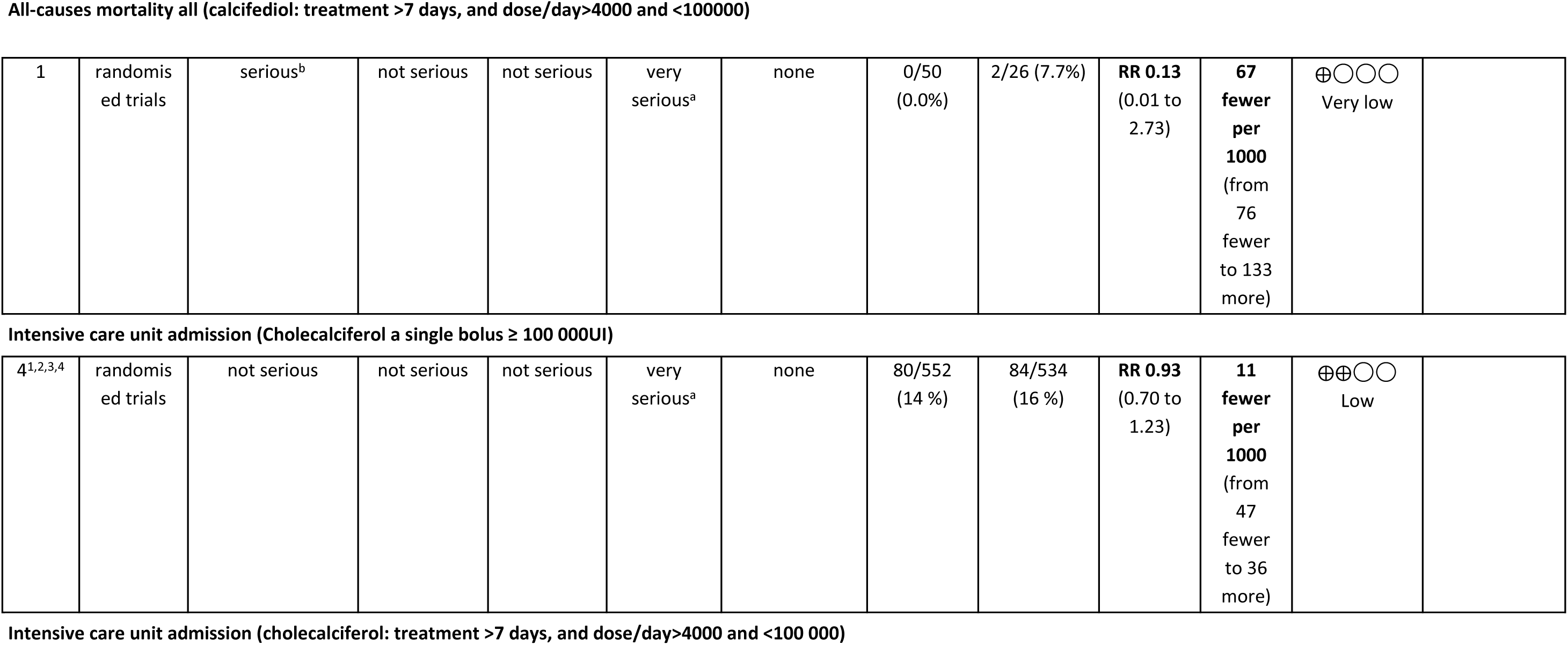
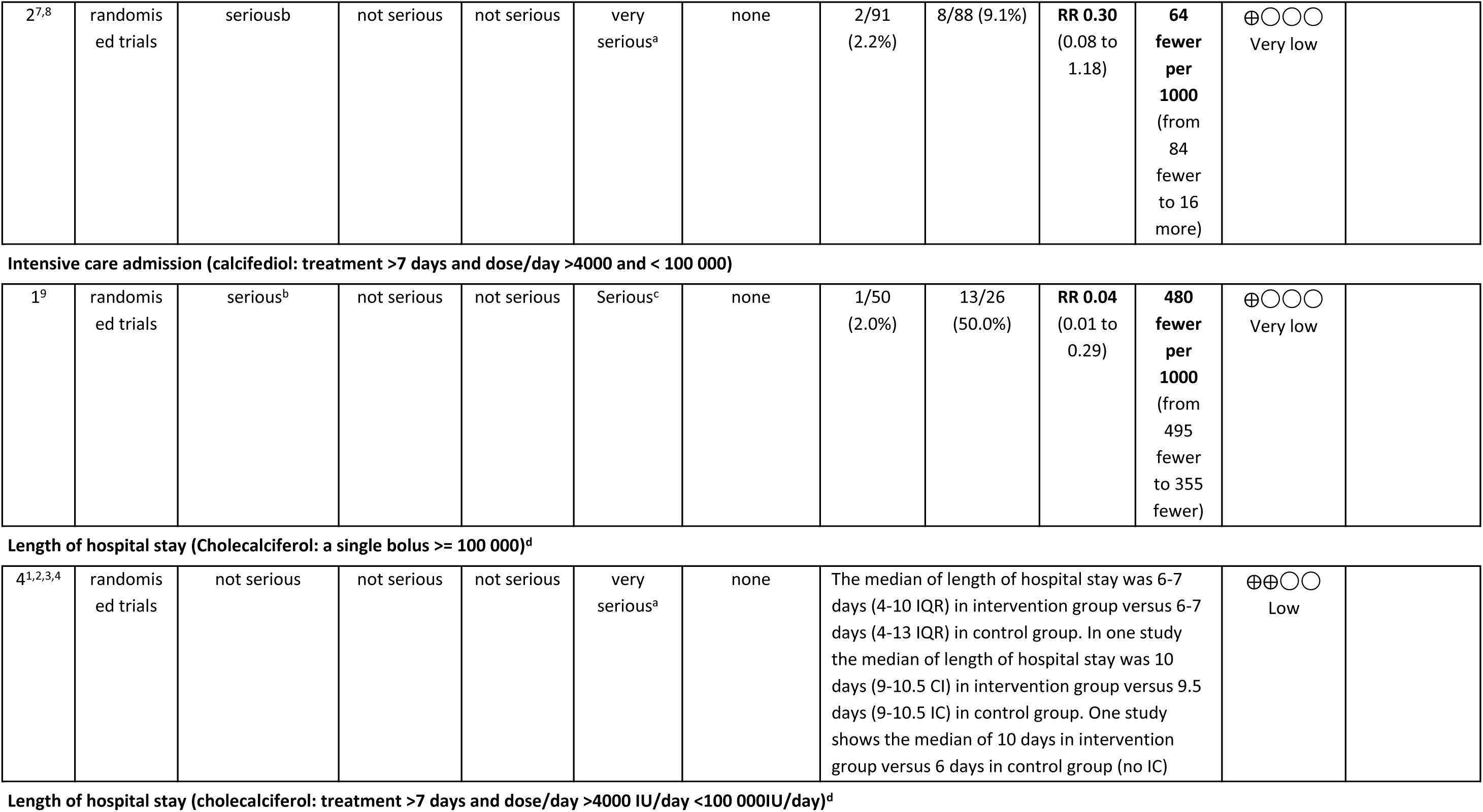
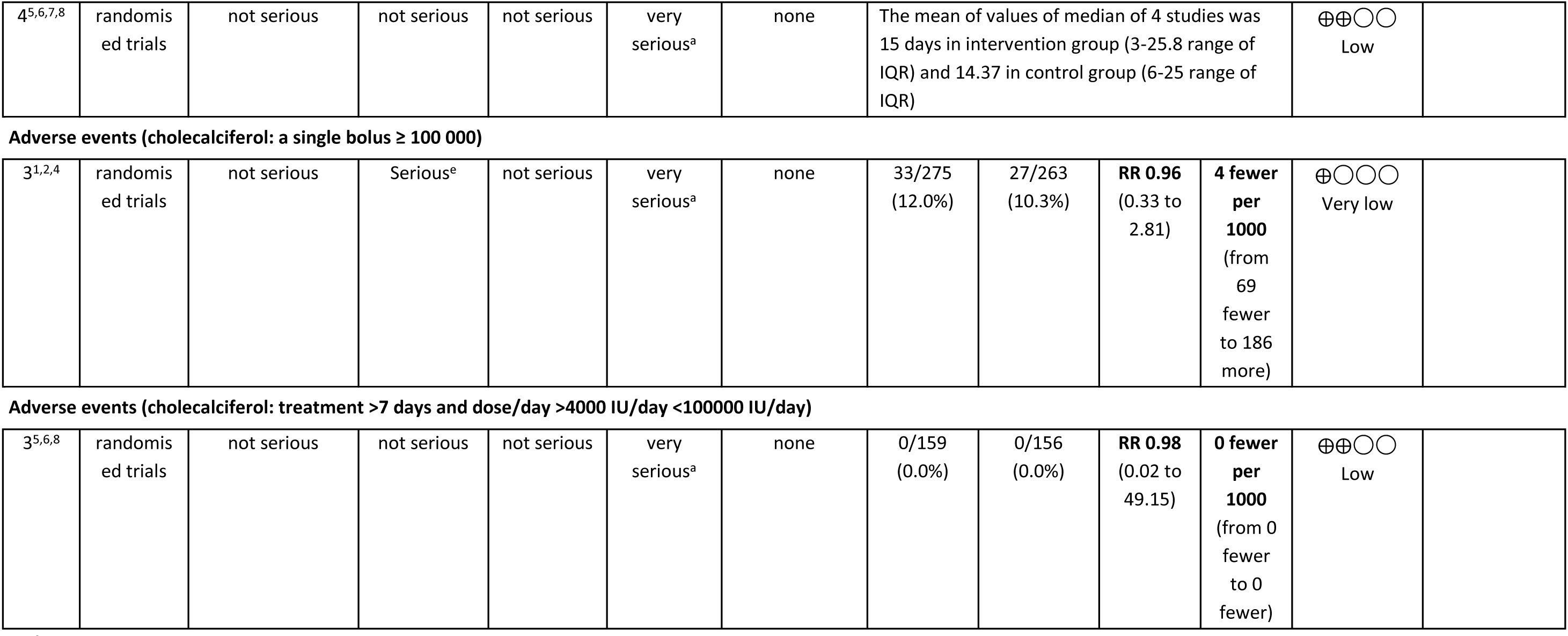
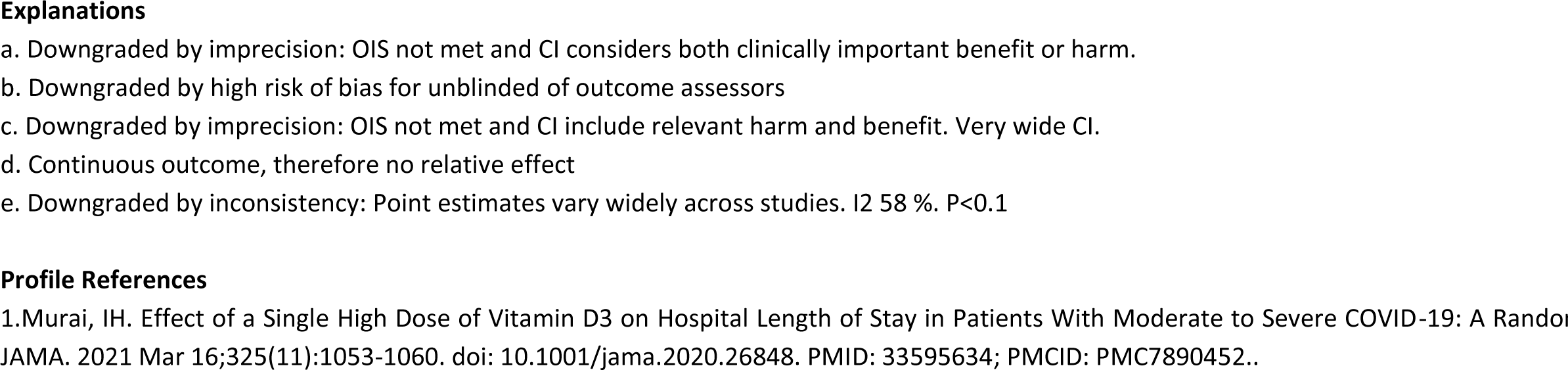
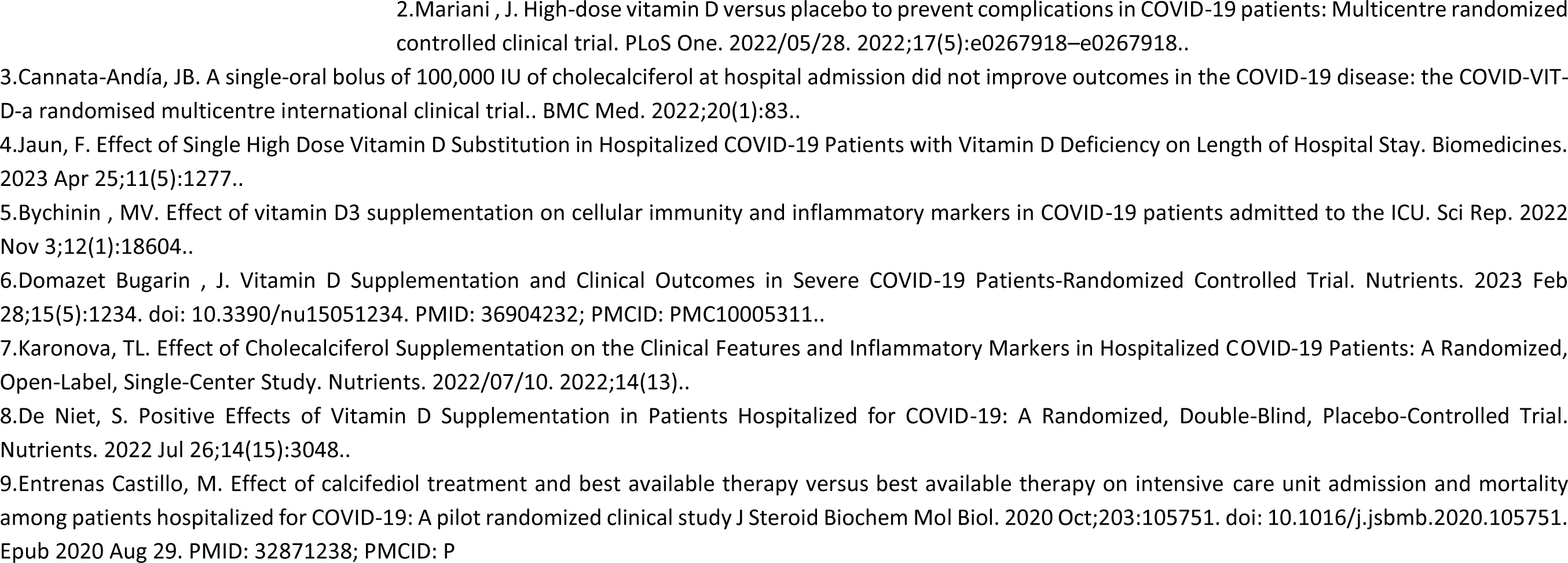

